# Nicotinamide riboside improves muscle mitochondrial biogenesis, satellite cell differentiation and gut microbiota composition in a twin study

**DOI:** 10.1101/2022.04.27.22274380

**Authors:** Helena Lapatto, Minna Kuusela, Aino Heikkinen, Maheswary Muniandy, Birgitta W. van der Kolk, Swetha Gopalakrishnan, Martin Sandvik, Mark S. Schmidt, Sini Heinonen, Sina Saari, Noora Pöllänen, Juho Kuula, Antti Hakkarainen, Janne Tampio, Tuure Saarinen, Marja-Riitta Taskinen, Nina Lundbom, Per-Henrik Groop, Marja Tiirola, Pekka Katajisto, Marko Lehtonen, Charles Brenner, Jaakko Kaprio, Satu Pekkala, Miina Ollikainen, Kirsi H. Pietiläinen, Eija Pirinen

**Affiliations:** Research Program for Clinical and Molecular Metabolism, Faculty of Medicine, University of Helsinki, FIN-00290 Helsinki, Finland; Institute for Molecular Medicine Finland (FIMM), HiLIFE, University of Helsinki, Helsinki, Finland; Institute of Biotechnology, HiLIFE, University of Helsinki, Helsinki, Finland; Department of Biochemistry, Carver College of Medicine, University of Iowa, Iowa City, Iowa 52242, USA; Department of Radiology, Medical Imaging Center, University of Helsinki and Helsinki University Hospital, Helsinki, Finland; School of Pharmacy, University of Eastern Finland, Kuopio, Finland; Department of Gastrointestinal Surgery, Helsinki University Hospital, Abdominal Center, Helsinki, Finland; Folkhälsan Institute of Genetics, Folkhälsan Research Center, Helsinki, Finland; Abdominal Center, Nephrology, University of Helsinki and Helsinki University Hospital, Helsinki, Finland; Department of Diabetes, Central Clinical School, Monash University, Melbourne, Australia; Department of Environmental and Biological Sciences, University of Jyväskylä, FI-40014 Jyväskylä, Finland; Department of Cell and Molecular Biology, Karolinska Institutet, Stockholm, Sweden; Department of Diabetes & Cancer Metabolism, City of Hope National Medical Center, Duarte, CA, USA; Faculty of Sport and Health Sciences, University of Jyväskylä, FI-40014 Jyväskylä, Finland; Obesity Research Unit, Research Program for Clinical and Molecular Metabolism, Faculty of Medicine, University of Helsinki, FIN-00290 Helsinki, Finland; Obesity Centre, Abdominal Centre, Endocrinology, Helsinki University Hospital and University of Helsinki, Helsinki, Finland; Research Unit for Internal Medicine, Faculty of Medicine, University of Oulu, FIN-90220 Oulu Finland

**Keywords:** Nicotinamide riboside, NAD^+^, vitamin B3, satellite cells, BMI, muscle, white adipose tissue, mitochondrial biogenesis, gut microbiota, twins

## Abstract

NAD^+^ precursor nicotinamide riboside (NR) has emerged as a promising compound to improve obesity-associated mitochondrial dysfunction and metabolic syndrome in mice. However, most short-term clinical trials conducted so far have failed to report positive outcomes. Therefore, we aimed to determine whether long-term NR supplementation boosts mitochondrial biogenesis and metabolic health in humans. Twins from 22 BMI-discordant monozygotic pairs were supplemented with an escalating dose of NR (250-1000 mg/day) for 5 months (clinicaltrials.gov entry NCT03951285). NR improved blood and tissue NAD^+^ metabolism, muscle mitochondrial number, myoblast differentiation and gut microbiota composition independent of BMI. NR also showed a capacity to modulate epigenetic control of gene expression in muscle and adipose tissue. However, NR did not ameliorate adiposity or metabolic health. Overall, our results suggest that NR acts as a potent modifier of NAD^+^ metabolism, muscle mitochondrial biogenesis and stem cell function, gut microbiota, and DNA methylation in humans irrespective of BMI.

## Introduction

Activation of mitochondria is an attractive treatment option for obesity and its related metabolic complications (Jokinen et al., 2017). Increasing intracellular levels of NAD^+^, the crucial cofactor for mitochondrial energy production, has shown great promise to improve mitochondrial number and oxidative capacity and to combat obesity and its related diseases in mice (Cantó et al., 2015). This observation has generated a high interest on whether mitochondrial dysfunction and metabolic health can be improved by NAD^+^ boosters in humans.

The intracellular NAD^+^ levels can be increased by supplementation with NAD^+^ precursors (Cantó et al., 2015). Vitamin B3 forms niacin (nicotinic acid), nicotinamide (NAM) and nicotinamide riboside (NR) are naturally occurring precursors of NAD^+^. They are converted to NAD^+^ via their distinct biosynthesis routes; Preiss-Handler and salvage pathways (Bogan and Brenner, 2008). The clinical use of niacin and NAM is challenged by dose-dependent adverse effects such as hepatotoxicity (Rajman et al., 2018). Niacin also induces cutaneous flushing through activation of G protein–coupled receptor 109A (Ganji et al., 2014). However, our previous study established that niacin can improve systemic NAD^+^ levels and muscle mitochondrial metabolism in humans without severe side-effects (Pirinen et al., 2020). The newest vitamin B3 family member NR (Bieganowski and Brenner, 2004), has been shown to increase muscle mitochondrial biogenesis and function, protect against diet-induced obesity and insulin resistance, and improve gut microbiota composition in mice (Canto et al., 2012; Cerutti et al., 2014; Frederick et al., 2016; Lozada-Fernandez et al., 2022; Trammell et al., 2016b; Yu et al., 2021; Zhang et al., 2016). Therefore, NR has rapidly entered clinical trials.

To this date, clinical studies have shown NR to be safe and to raise NAD^+^ levels in whole blood, peripheral blood mononuclear cells, urine, muscle and brain with a dose from 300 mg/day up to 2000 mg/day (Airhart et al., 2017; Brakedal et al., 2022; Dollerup et al., 2018; Elhassan et al., 2019; Martens et al., 2018; Remie et al., 2020; Trammell et al., 2016a). Unfortunately, most human trials have failed to show significant improvements in adiposity and metabolic health in healthy overweight or obese individuals after 3-to-12-week NR supplementation (Airhart et al., 2017; Dollerup et al., 2018; Elhassan et al., 2019; Martens et al., 2018; Remie et al., 2020; Trammell et al., 2016a). However, improved body composition, physical performance, muscle acetylcarnitine levels, arterial stiffness and blood pressure or decreased circulating inflammatory cytokines have been reported in a few studies with a short-term follow-up (Dolopikou et al., 2020; Elhassan et al., 2019; Remie et al., 2020). At present, the evidence of the efficacy of NR on tissue mitochondrial biogenesis and function still relies on data on mice as positive outcomes are lacking from clinical studies (Dollerup et al., 2020; Remie et al., 2020). Also, as the number of human intervention studies with NR is still relatively low and the longest clinical trial conducted so far has been 12 weeks, interventions of a longer duration are required.

We hypothesized that long-term NR supplementation could improve mitochondrial biogenesis in humans. Therefore, our primary outcome was the change in mitochondrial biogenesis. We supplemented twins from body mass index (BMI)-discordant monozygotic (MZ) twin pairs with NR for 5 months and investigated the effect of NR on tissue mitochondrial biogenesis but also on metabolic health. As MZ twins in a pair have virtually the same genomic sequence, this allowed us to investigate whether BMI is a key factor in determining the response to NR. Additionally, we compared the effect of NR to placebo in a small cohort of BMI-concordant MZ twin pairs where one of the twins in each pair received placebo while the other twin received NR. We found that 5-month NR supplementation improves muscle mitochondrial biogenesis, muscle myoblast differentiation and microbiota composition, and modulates DNA methylation, with potential effects on epigenetic control of gene expression, in both twins from the BMI-discordant pairs, *i.e.* regardless of BMI.

However, these changes were not translated into improvements in adiposity or metabolic status. In fact, there was an overall increase in body weight and a decrease in insulin sensitivity during the study. Together, our study underscores that long-term NR supplementation can affect various metabolic processes in humans.

## Results

### Characteristics of study participants

Altogether 22 BMI-discordant and four BMI-concordant MZ twin pairs, aged ~40 (interquartile range 33 - 41), 44% females, participated in the study (Table S1). All twins from the BMI-discordant pairs were supplemented with NR. Of the concordant twin pairs (Table S1), one cotwin was randomized to placebo and the other one to NR. The daily NR dose was gradually escalated by 250 mg/week, to the full dose 1000 mg/day, continuing until 5 months (Figure 1A). The primary endpoint was the change in muscle mitochondrial biogenesis (clinicaltrials.gov entry NCT03951285). For the list of secondary endpoints, see https://clinicaltrials.gov/ct2/show/NCT03951285?term=nicotinamide+riboside&draw=4&rank=29. Figure 1A and Figure S1 present the study design and the procedures for the selection of study subjects and data analyses, respectively.

**Figure 1.**
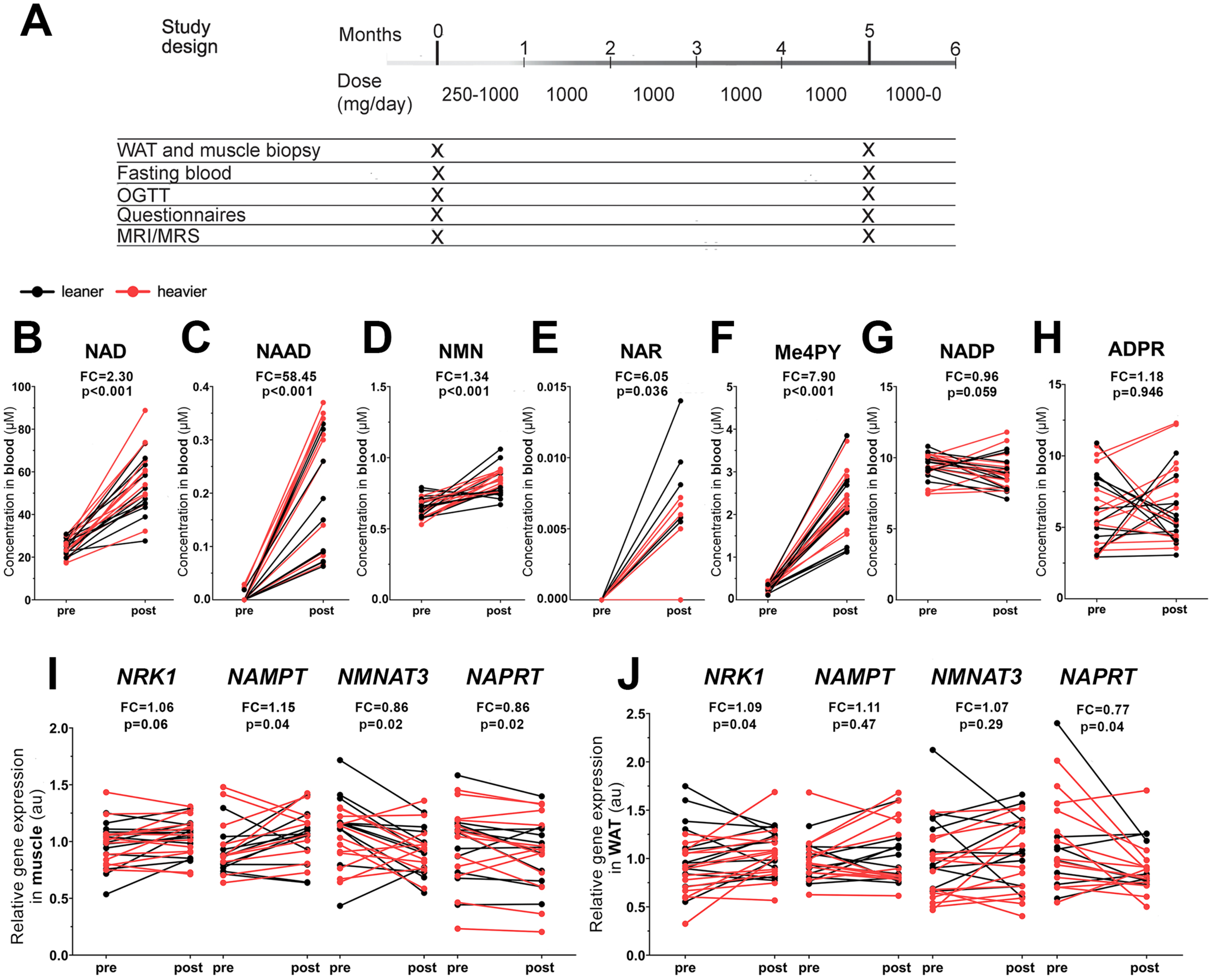
NR increases whole blood NAD^+^ levels and tissue NAD^+^ biosynthesis in the twins from the BMI-discordant pairs. **(A)** The study protocol. The daily NR dose was gradually escalated from 250 mg/day by 250 mg per every week to achieve the final treatment dose, 1 g/day. At the end of the study, the dose was decreased by 250 mg per every week. Clinical examinations and collection of fasting blood samples, muscle and WAT biopsies were performed at baseline and after 5 months. **(B-H)** Whole blood NAD^+^ metabolite levels pre versus post NR (n=14 twin pairs/28 individuals). **(I-J)** Expression of genes involved in NAD^+^ biosynthesis in muscle (I) and WAT (J) pre versus post NR (n=10-13 twin pairs/21-26 individuals). Arbitrary unit (au) indicates the relative target gene expression normalized to the expression of reference genes. Pre and post values of all individuals are connected with a line; leaner cotwins are presented with black color and heavier cotwins of the BMI-discordant pairs with red color. Fold change (FC) indicates the mean of post NR value divided by the pre NR value. Paired Wilcoxon signed-rank test was used as a statistical test to determine the effect of NR in all twins from the BMI-discordant pairs. NAR, nicotinic acid riboside; ADPR, ADP ribose. See also Figures S1-S2 and Table S2.

The heavier cotwins of the BMI-discordant twin pairs (BMI mean 33 ± SD 6) had expectedly higher measures of adiposity, insulin resistance, basal metabolic rate, liver fat and enzyme alanine aminotransferase (ALT) than their leaner cotwins (BMI 28 ± 4) (Table S1). The total intake of food or macronutrients, or physical activity did not differ between the leaner and heavier cotwins of the BMI-discordant pairs, but the heavier cotwins consumed more alcohol (Table S1). Both twins from the BMI-concordant pairs randomly assigned to receive either NR (BMI 32 ± 7) or placebo (BMI 32 ± 7) appeared very similar in their baseline characteristics (Table S1). Overall, most of the clinical values were within reference values in both twin pair groups (Table S1).

### NR increases whole blood NAD^+^ concentrations and tissue NAD^+^ biosynthesis

To elucidate the compliance and the effect of NR on NAD^+^ metabolism in the twins from the BMI-discordant pairs, we analyzed NAD metabolome by targeted liquid chromatography–mass spectrometry on the whole blood samples. At baseline, blood NAD metabolites did not differ between the heavier or leaner cotwins (Figures S2A-S2F). NR boosted whole blood NAD^+^ levels by 2.3-fold (Figure 1B) in all twins from the BMI-discordant pairs. In line, nicotinic acid adenine dinucleotide (NAAD), the validated biomarker for NR supplementation and an enhanced rate of NAD^+^ synthesis (Trammell et al., 2016a), was elevated upon NR (Figure 1C). NR also significantly increased the levels of nicotinamide mononucleotide (NMN), the phosphorylated form of NR, (Figure 1D) and nicotinic acid riboside, the deamidated form of NR (Figure 1E). N-methyl-4-pyridone-5-carboxamide (Me4py), was elevated by 8-fold (Figure 1F) suggesting an enhanced elimination of NR’s degradation product NAM via methylation. Of the other NAD metabolites, NADP trended to decrease (Figure 1G) while ADP ribose did not significantly change (Figure 1H). BMI did not affect the response to NR (the delta changes, *i.e.* the changes from baseline to 5 months), except that the increase in Me4Py levels was more pronounced in the heavier cotwins compared with the leaner cotwins from the BMI discordant pairs (Table S2). Overall, NR was effectively metabolized, and it had a great potency to increase blood NAD metabolites.

To investigate the influence of NR on tissue NAD^+^ biosynthesis, we measured mRNA expression of NAD^+^ biosynthetic enzymes from muscle and white adipose tissue (WAT). At baseline, in comparison to the leaner cotwins of the BMI-discordant pairs, the heavier cotwins exhibited significantly lower expression of *NMNAT3,* one of the *NAMN*/*NMN* adenylyltransferase isoforms, in muscle and WAT (Figures S2G-S2H). The expression of the NR metabolizing enzyme, nicotinamide riboside kinase 1 (*NRK1*), was also lower in the WAT of the heavier cotwins (Figures S2G-S2H). In the twins from the BMI-discordant pairs, NR increased the muscle expression of NAM phosphoribosyltransferase (*NAMPT)*, the enzyme converting NAM towards NAD^+^, and trended to upregulate *NRK1* while decreasing *NMNAT3* (Figure 1I). In WAT, *NRK1* expression was significantly elevated while *NAMPT* and *NMNAT3* were unchanged upon NR (Figure 1J). Interestingly, NR downregulated the Preiss-Handler pathway enzyme nicotinic acid phosphoribosyltransferase (NAPRT) both in muscle and WAT (Figures 1I-1J) indicating decreased niacin utilization and/or increased reliance on the salvage pathway enzymes upon NR. The NR-induced alterations in the mRNA expression of NAD^+^ biosynthetic enzymes were similar in both twins from the BMI discordant pairs, except that NR tended to increase WAT *NRK1* expression more in the heavier cotwins compared with the leaner cotwins (Table S2). As a whole, NR promoted both muscle and WAT NAD^+^ biosynthesis via the salvage pathways regardless of BMI.

### Body composition and lifestyle factors

We next determined the impact of NR on body composition in the twins from the BMI-discordant pairs. During the study, body weight (~3 kg) and the whole-body fat percentage increased significantly, and the size of all adipose tissue depots tended to elevate in all twins from the BMI discordant pairs (Table 1). There were no significant changes in adipocyte number, diameter, volume and weight upon NR (Table 1). However, the expression of peroxisome proliferator-activated receptor γ (*PPAR*γ), the essential transcription factor controlling adipogenesis, was upregulated after NR treatment (Table 1). Lean tissue (muscle) and bone mass, and liver fat content remained unaltered (Table 1). Basal metabolic rate significantly enhanced probably due to the increase in body weight (Table 1). Food diaries and questionnaires showed no marked changes in the intake of food, macronutrients, niacin equivalent, alcohol or physical activity (Table 1). The changes from baseline to 5 months did not differ between the leaner and heavier cotwins (Table S2). Altogether, body weight and the fat percentage increased in the twins from the BMI-discordant pairs during the intervention, but we did not observe clear changes in the self-reported lifestyle measures.

**Table 1.**
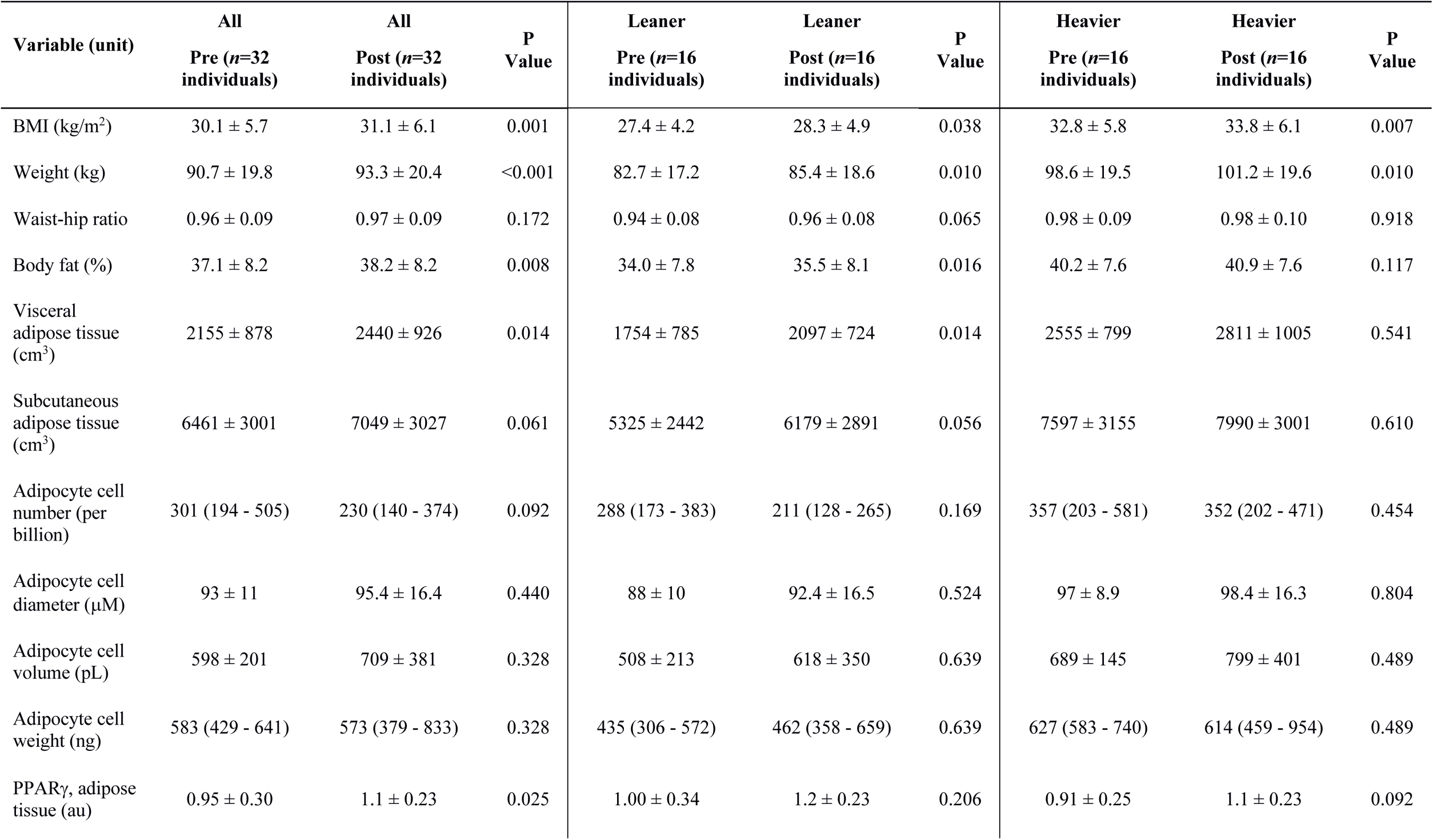
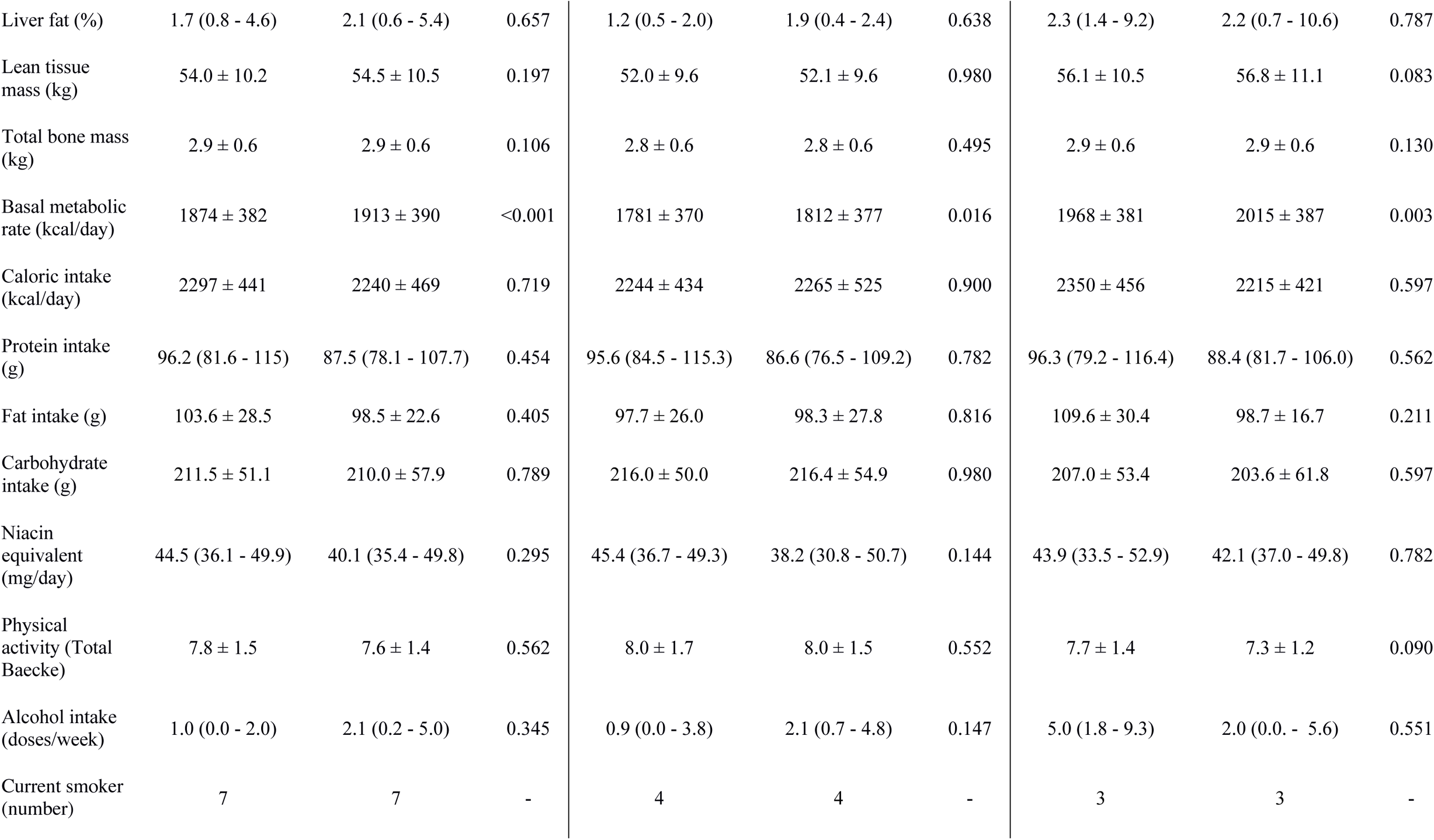
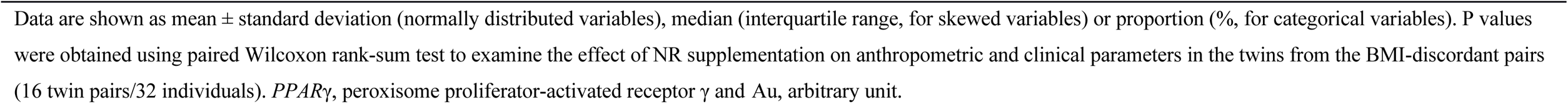
The effect of NR on body composition and lifestyle factors in the twins from the BMI-discordant pairs.

| Variable (unit) | All<br>Pre (*n*=32<br>individuals) | All<br>Post (*n*=32<br>individuals) | P<br>Value | Leaner<br>Pre (*n*=16<br>individuals) | Leaner<br>Post (*n*=16<br>individuals) | P<br>Value | Heavier<br>Pre (*n*=16<br>individuals) | Heavier<br>Post (*n*=16<br>individuals) | P<br>Value |
| --- | --- | --- | --- | --- | --- | --- | --- | --- | --- |
| BMI (kg/m<sup>2</sup>) | 30.1 ± 5.7 | 31.1 ± 6.1 | 0.001 | 27.4 ± 4.2 | 28.3 ± 4.9 | 0.038 | 32.8 ± 5.8 | 33.8 ± 6.1 | 0.007 |
| Weight (kg) | 90.7 ± 19.8 | 93.3 ± 20.4 | <0.001 | 82.7 ± 17.2 | 85.4 ± 18.6 | 0.010 | 98.6 ± 19.5 | 101.2 ± 19.6 | 0.010 |
| Waist-hip ratio | 0.96 ± 0.09 | 0.97 ± 0.09 | 0.172 | 0.94 ± 0.08 | 0.96 ± 0.08 | 0.065 | 0.98 ± 0.09 | 0.98 ± 0.10 | 0.918 |
| Body fat (%) | 37.1 ± 8.2 | 38.2 ± 8.2 | 0.008 | 34.0 ± 7.8 | 35.5 ± 8.1 | 0.016 | 40.2 ± 7.6 | 40.9 ± 7.6 | 0.117 |
| Visceral<br>adipose tissue<br>(cm<sup>3</sup>) | 2155 ± 878 | 2440 ± 926 | 0.014 | 1754 ± 785 | 2097 ± 724 | 0.014 | 2555 ± 799 | 2811 ± 1005 | 0.541 |
| Subcutaneous<br>adipose tissue<br>(cm<sup>3</sup>) | 6461 ± 3001 | 7049 ± 3027 | 0.061 | 5325 ± 2442 | 6179 ± 2891 | 0.056 | 7597 ± 3155 | 7990 ± 3001 | 0.610 |
| Adipocyte cell<br>number (per<br>billion) | 301 (194 - 505) | 230 (140 - 374) | 0.092 | 288 (173 - 383) | 211 (128 - 265) | 0.169 | 357 (203 - 581) | 352 (202 - 471) | 0.454 |
| Adipocyte cell<br>diameter (μM) | 93 ± 11 | 95.4 ± 16.4 | 0.440 | 88 ± 10 | 92.4 ± 16.5 | 0.524 | 97 ± 8.9 | 98.4 ± 16.3 | 0.804 |
| Adipocyte cell<br>volume (pL) | 598 ± 201 | 709 ± 381 | 0.328 | 508 ± 213 | 618 ± 350 | 0.639 | 689 ± 145 | 799 ± 401 | 0.489 |
| Adipocyte cell<br>weight (ng) | 583 (429 - 641) | 573 (379 - 833) | 0.328 | 435 (306 - 572) | 462 (358 - 659) | 0.639 | 627 (583 - 740) | 614 (459 - 954) | 0.489 |
| PPARγ, adipose<br>tissue (au) | 0.95 ± 0.30 | 1.1 ± 0.23 | 0.025 | 1.00 ± 0.34 | 1.2 ± 0.23 | 0.206 | 0.91 ± 0.25 | 1.1 ± 0.23 | 0.092 |
| Liver fat (%) | 1.7 (0.8 - 4.6) | 2.1 (0.6 - 5.4) | 0.657 | 1.2 (0.5 - 2.0) | 1.9 (0.4 - 2.4) | 0.638 | 2.3 (1.4 - 9.2) | 2.2 (0.7 - 10.6) | 0.787 |
| Lean tissue mass (kg) | 54.0 ± 10.2 | 54.5 ± 10.5 | 0.197 | 52.0 ± 9.6 | 52.1 ± 9.6 | 0.980 | 56.1 ± 10.5 | 56.8 ± 11.1 | 0.083 |
| Total bone mass (kg) | 2.9 ± 0.6 | 2.9 ± 0.6 | 0.106 | 2.8 ± 0.6 | 2.8 ± 0.6 | 0.495 | 2.9 ± 0.6 | 2.9 ± 0.6 | 0.130 |
| Basal metabolic rate (kcal/day) | 1874 ± 382 | 1913 ± 390 | <0.001 | 1781 ± 370 | 1812 ± 377 | 0.016 | 1968 ± 381 | 2015 ± 387 | 0.003 |
| Caloric intake (kcal/day) | 2297 ± 441 | 2240 ± 469 | 0.719 | 2244 ± 434 | 2265 ± 525 | 0.900 | 2350 ± 456 | 2215 ± 421 | 0.597 |
| Protein intake (g) | 96.2 (81.6 - 115) | 87.5 (78.1 - 107.7) | 0.454 | 95.6 (84.5 - 115.3) | 86.6 (76.5 - 109.2) | 0.782 | 96.3 (79.2 - 116.4) | 88.4 (81.7 - 106.0) | 0.562 |
| Fat intake (g) | 103.6 ± 28.5 | 98.5 ± 22.6 | 0.405 | 97.7 ± 26.0 | 98.3 ± 27.8 | 0.816 | 109.6 ± 30.4 | 98.7 ± 16.7 | 0.211 |
| Carbohydrate intake (g) | 211.5 ± 51.1 | 210.0 ± 57.9 | 0.789 | 216.0 ± 50.0 | 216.4 ± 54.9 | 0.980 | 207.0 ± 53.4 | 203.6 ± 61.8 | 0.597 |
| Niacin equivalent (mg/day) | 44.5 (36.1 - 49.9) | 40.1 (35.4 - 49.8) | 0.295 | 45.4 (36.7 - 49.3) | 38.2 (30.8 - 50.7) | 0.144 | 43.9 (33.5 - 52.9) | 42.1 (37.0 - 49.8) | 0.782 |
| Physical activity (Total Baecke) | 7.8 ± 1.5 | 7.6 ± 1.4 | 0.562 | 8.0 ± 1.7 | 8.0 ± 1.5 | 0.552 | 7.7 ± 1.4 | 7.3 ± 1.2 | 0.090 |
| Alcohol intake (doses/week) | 1.0 (0.0 - 2.0) | 2.1 (0.2 - 5.0) | 0.345 | 0.9 (0.0 - 3.8) | 2.1 (0.7 - 4.8) | 0.147 | 5.0 (1.8 - 9.3) | 2.0 (0.0 - 5.6) | 0.551 |
| Current smoker (number) | 7 | 7 | - | 4 | 4 | - | 3 | 3 | - |

Data are shown as mean ± standard deviation (normally distributed variables), median (interquartile range, for skewed variables) or proportion (% for categorical variables). P values were obtained using paired Wilcoxon rank-sum test to examine the effect of NR supplementation on anthropometric and clinical parameters in the twins from the BMI-discordant pairs (16 twin pairs/32 individuals). *PPAR* $\gamma$ , peroxisome proliferator-activated receptor $\gamma$ and Au, arbitrary unit.

### Insulin sensitivity and cardiovascular health

To understand whether the increased body weight resulted in alterations in glucose homeostasis in the twins from the BMI-discordant pairs, we ran a comprehensive set of measures during an oral glucose tolerance test (OGTT). Glucose, insulin, C-peptide, HOMA and Matsuda index indicated impairments in all twins from the BMI-discordant pairs upon NR, but values mostly remained within the reference ranges (Table S3). No significant differences were observed regarding the changes from baseline to 5 months between the heavier and leaner cotwins from the BMI discordant pairs, except that HbA1c increased slightly more in the leaner cotwins compared with the heavier cotwins (Table S2). Overall, our data suggest that insulin sensitivity decreased during the study.

Next, we elucidated whether cardiovascular health was affected by NR. Except for the small, clinically nonsignificant decrease in HDL and triglycerides, no changes in blood lipid values were detected in the twins from the BMI-discordant pairs upon NR (Table S4). Neither were there any changes in blood pressure, pulse rate, nor in the inflammation marker, high-sensitivity C-reactive protein (CRP) (Table S4). BMI did not influence NR’s effect on these analyzed variables (Table S2).

### NR boosts mitochondrial biogenesis in muscle but not in WAT

Given that NR significantly elevated whole-blood and tissue NAD^+^ biosynthesis in the twins from the BMI-discordant pairs, we determined the effect of NR on our primary outcome, muscle mitochondrial biogenesis, in these twins. Transmission electron micrographs (TEM) of muscle samples showed that mitochondria became more abundant (~14%) in the intermyofibrillar region of type I muscle fibers upon NR (Figures 2A-2B). In addition, mitochondria covered a larger cross-sectional area of the muscle fiber area after NR supplementation (Figure 2C). Morphological analysis of mitochondria (perimeter, diameter, form *i.e.* branching factor, or aspect *i.e.* length-to-width ratio) did not reveal any alterations upon NR (Figures 2D-2G). In line with TEM findings, NR clearly elevated (~30%) muscle mtDNA amount (Figure 2H) and muscle expression of the following transcription factors regulating mitochondrial biogenesis; estrogen-related receptor α *(ERRα*), transcription factor A *(TFAM)* and mitofusin 2 (*MFN2)* (Figure 2I). In addition, NR significantly elevated the expression of muscle OXPHOS complex subunits cytochrome c oxidase subunit 4 (*COX4)* and ATP synthase *α* (*ATP5A)* but downregulated one complex I subunit (Figures S3A-S3B). The effect of NR on muscle mitochondrial number, mtDNA amount, transcription factors and OXPHOS complex subunits’ expression was not dependent on BMI (Table S2). Collectively, NR promoted muscle biogenesis likely via *ESRRA/TFAM/MFN2*.

**Figure 2.**
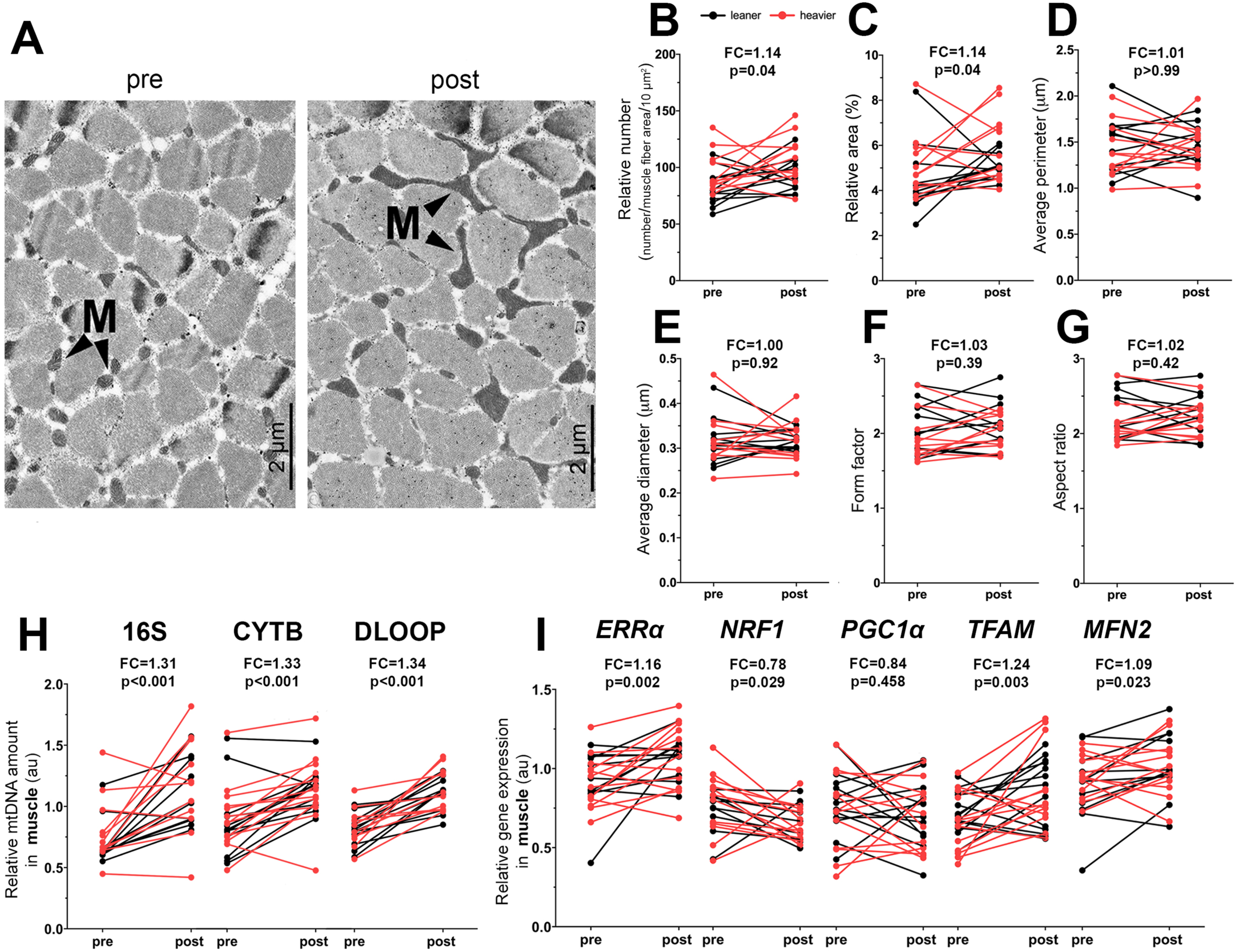
NR boosts muscle mitochondrial biogenesis in the twins from the BMI-discordant pairs. **(A)** TEM images of the muscle intermyofibrillar mitochondria (M) in one representative study participant pre and post NR. 2000x magnification. Scale bar 2 μm. **(B-G)** Quantification of (B) the number of mitochondria per 10 µm^2^ relative to the muscle fiber area, (C) percentage of mitochondrial surface area per total muscle fiber area, (D) an average perimeter of mitochondria, (E) an average diameter of mitochondria, (F) a form factor (the branching of mitochondria), (G) an aspect ratio (the length-to-width ratio of mitochondria) in the muscle biopsies (n=11 twin pairs/22 individuals). **(H)** Muscle relative mtDNA amount presented as a ratio of mtDNA genome (16S, CYTB and DLOOP) per nuclear genome pre versus post NR (n=11-12 twin pairs/22-25 individuals). 16S, 16 rRNA; CYTB, cytochrome b and DLOOP, D-loop region. **(I)** Gene expression of the key regulators of mitochondrial biogenesis in muscle pre versus post NR (n=10-12 twin pairs/21-28 individuals). Arbitrary unit (au) indicates the relative gene expression normalized to the expression of reference genes. Pre and post values of all individuals are connected with a line; leaner cotwins are presented with black color and heavier cotwins of the BMI-discordant pairs with red color. Fold change (FC) indicates the mean of post NR value divided by the pre NR value. Paired Wilcoxon signed-rank test was used as a statistical test to determine the effect of the treatment in all twins from the BMI-discordant pairs. See also Figure S3 and Table S2.

We next investigated the effect of NR on WAT mitochondria in the twins from the BMI-discordant pairs. NR did not significantly influence mtDNA amount and had only a minor effect on the gene expression profile in WAT irrespective of BMI (Figures S3C-S3F, and Table S2).

### NR promotes muscle satellite cell differentiation

As NR has been previously shown to increase the number and the function of muscle stem cells *i.e*., satellite cells in mice (Zhang et al., 2016), we investigated the effect of NR on muscle satellite cells in the twins from the BMI-discordant pairs. Interestingly, muscle expression of the stem cell marker, paired box 7 (*PAX7*), and the number of PAX7^+^ satellite cells were significantly reduced upon NR (Figures 3A-3C). To characterize the satellite cells, we created primary myoblast cultures from the muscle biopsies. Myoblasts from the NR-treated twins showed reduced stemness and increased differentiation, as demonstrated by a significant decrease in the ratio of *PAX7* and the myogenic marker myogenin *(MYOG)* (Figure 3D). BMI did not influence the observed changes in *PAX7* expression, satellite cell numbers or *PAX7*/*MYOG* ratio upon NR (Table S2). Thus, our data indicate that NR likely facilitated the activation of satellite cells towards differentiation instead of self-renewal. *In vivo* in the absence of injury, this may lead to the fusion of satellite cells to existing muscle fibers resulting in the observed decline in the satellite cell number.

**Figure 3.**
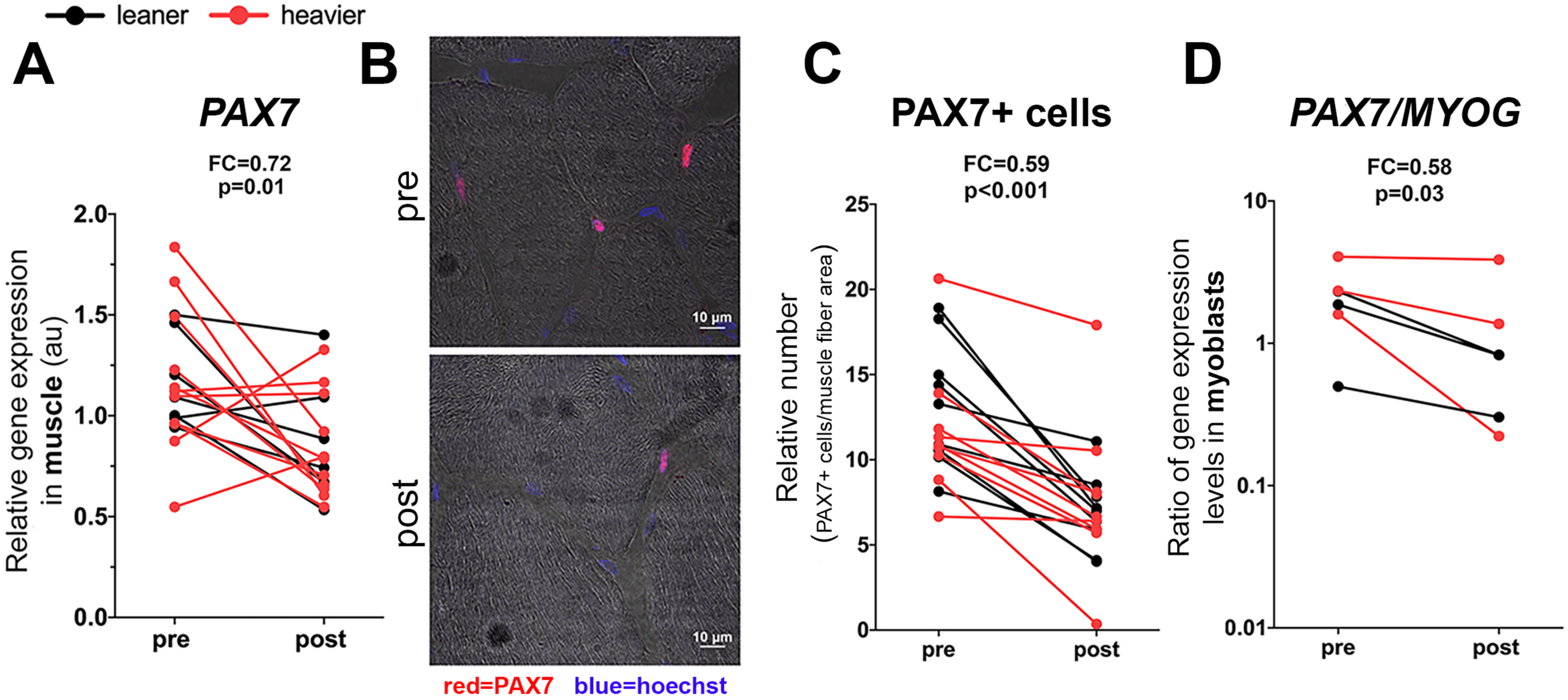
NR promotes muscle satellite cell differentiation in the twins from the BMI-discordant pairs. **(A)** Muscle gene expression level of satellite cell marker *PAX7* pre versus post NR (n=9 twin pairs/18 individuals). **(B)** Immunostaining of PAX7+ satellite cells in muscle cryosections pre versus post NR in one representative study participant. PAX7 (red=satellite cells); Hoechst (blue=nuclei). Scale bar 10 µm. **(C)** Muscle PAX7+ satellite cell quantification pre versus post NR (n=10 twin pairs/20 individuals). **(D)** Ratio of *PAX7/MYOG* mRNA expression in myoblasts pre versus post NR (n=3 twin pairs/6 individuals). Y-axis is on a logarithmic scale. Pre and post values of all individuals are connected with a line; leaner cotwins are presented with black color and heavier cotwins of the BMI-discordant pairs with red color. Arbitrary unit (au) indicates the relative target gene expression normalized to the expression of reference genes. Fold change (FC) indicates the mean of post NR value divided by the pre NR value. Paired Wilcoxon signed-rank test was used as a statistical test to determine the effect of the treatment on all cotwins from the BMI-discordant pairs. See also Table S2.

### NR may affect the epigenetic control of gene expression in muscle and WAT

The supplementation with NAD^+^ precursors has been suggested to induce a decline in methyl groups due to enhanced elimination of NAM via methylation (Sun et al., 2017). Thus, we measured the levels of circulating homocysteine, increased levels of which can be considered as a marker for compromised cellular methylation status. NR slightly but significantly increased the levels of total plasma homocysteine, especially in the leaner cotwins from the BMI discordant pairs (Figure 4A and Tables S2 and S4), but not beyond the normal range, suggesting a reduction in the cellular methylation capacity. These findings prompted us to investigate whether NR affected tissue DNA methylation. NR significantly reduced global DNA methylation level in muscle, but not in WAT (Figures 4B-4C), with similar effects in both cotwins irrespective of BMI (median within-pair difference 0.001, p=0.47 in muscle, and −0.001, p=0.52 in WAT) upon NR. Overall, our results suggest that long-term NR supplementation influences global DNA methylation in a tissue-specific manner.

**Figure 4.**
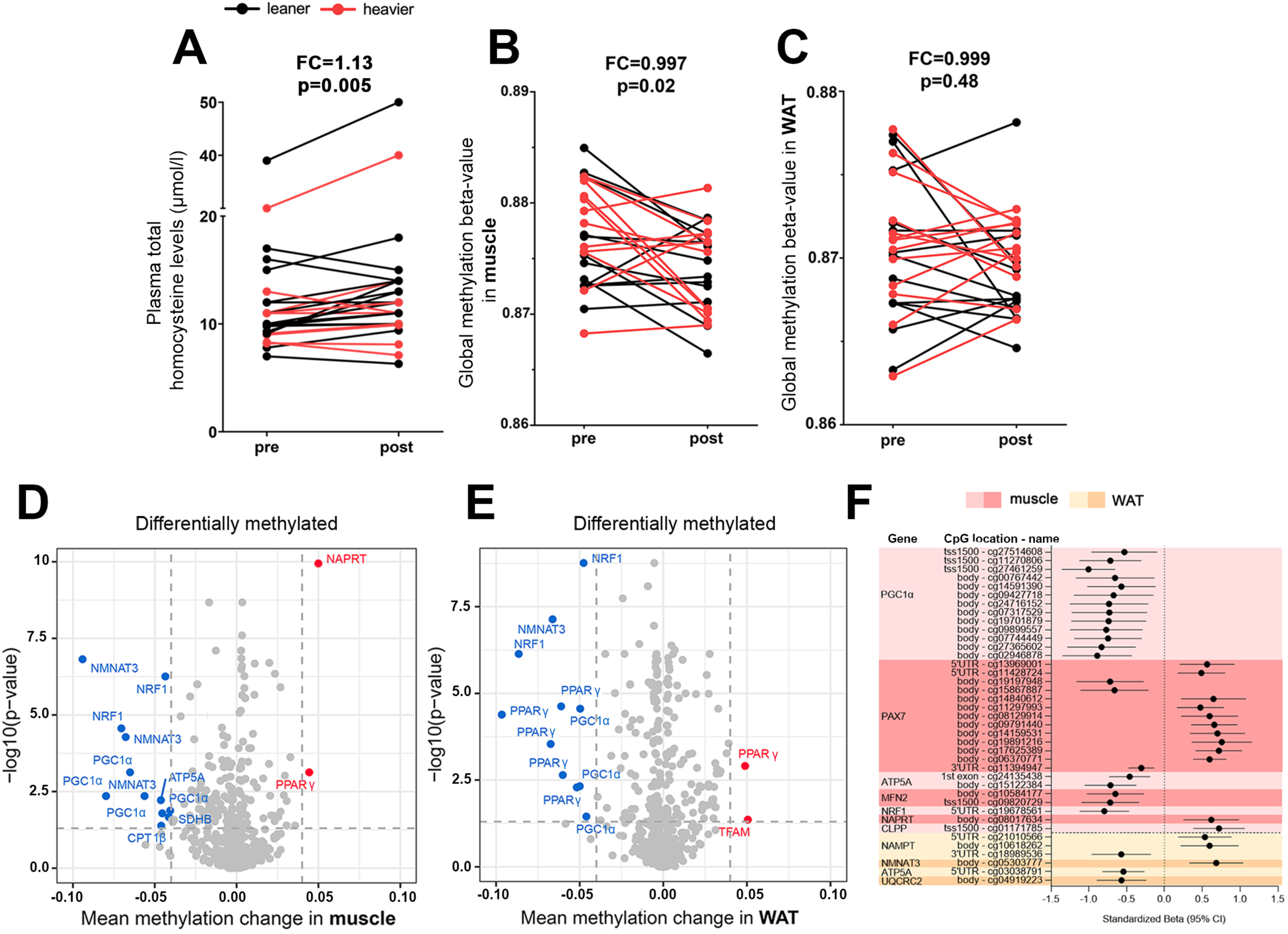
NR modifies the epigenetic control of gene expression in the twins from the BMI-discordant pairs. **(A)** Plasma total homocysteine levels pre versus post NR (n=13 twin pairs/26 individuals). **(B-C)** Global DNA methylation levels pre versus post NR in (B) muscle (n=12 twin pairs/24 individuals) and (C) WAT (n=13 twin pairs/26 individuals). Pre and post values of all individuals are connected with a line; leaner cotwins are presented with black color and heavier cotwins of the BMI-discordant pairs with red color. Fold change (FC) indicates the mean of post NR value divided by the pre NR value. **(D-E)** Volcano plots showing statistical significance (y-axis) and the magnitude of the change in mean methylation beta-value (x-axis) upon NR for each CpG site in (D) muscle (n=12 twin pairs/25 individuals) and (E) WAT (n=14 twin pairs/28 individuals). Each dot represents a single CpG site (n=619 and 518 in muscle and WAT, respectively). Highlighted are CpGs with FDR<0.05 (dashed horizontal line) and the mean methylation change (dashed vertical lines) larger than 0.04 (red dots) or smaller than −0.04 (blue dots). CpGs that do not meet the criteria are marked as grey. **(F)** Associations between CpG site methylation and gene expression in muscle (n=7 twin pairs/14 individuals) and WAT (n=9 twin pairs/18 individuals) upon NR. The red color indicates associations in muscle while yellow indicates associations in WAT. Error bars denote 95% confidence intervals. Only significant (FDR< 0.05) associations are shown. Standardized betas show how many standard deviations in methylation level changes are associated with one standard deviation of change in gene expression. Paired Wilcoxon signed-rank test was used as a statistical test to determine the effect of the NR supplementation on plasma total homocysteine in all twins from the BMI-discordant pairs. Statistical analyses for methylation data are described under the Method section See also Tables S2 and S5-S6.

Next, we performed differential DNA methylation analysis for selected genes involved in NAD^+^ biosynthesis, mitochondrial biogenesis, protein quality control and fatty acid oxidation, OXPHOS and satellite cell identity in pre versus post NR samples of the twins from the BMI discordant pairs. This was to identify CpG sites that show significantly different methylation levels in tissues upon NR. NR altered the methylation of 173/619 (28 %) and 210/518 (41 %) CpG sites in muscle and WAT, respectively (Figures 4D-4E). While NR induced both hypo- and hypermethylation, most of the analyzed CpG sites were hypomethylated (~60% in both tissues) in post-NR tissues. The hypomethylated CpGs were typically located at the open sea, whereas the hypermethylated sites were mostly observed at CpG islands. The majority of the hypomethylated CpGs were detected in *PPAR*γ coactivator 1α (*PGC-1α), NMNAT3 and* nuclear respiratory factor 1 *(NRF1)* in muscle and *PPAR*γ and *NRF1* in WAT. The heavier and leaner cotwins from the BMI discordant pairs did not differ in their response to NR at their tissue CpG site methylation (Table S5).

As DNA methylation may influence gene expression, we assessed associations between gene expression and CpG methylation of the respective genes. In muscle, a total of 33 statistically significant associations were observed (Table S6). All 13 associations observed between *PGC-1α* expression and methylation were negative (Figure 4F) suggesting that hypomethylation of *PGC-1α* may increase its transcription in muscle. In addition, 13 significant, mostly positive associations were detected between expression and methylation of *PAX7*. Thus, hypomethylation of *PAX7* may repress its expression in muscle. Other detected associations between gene expression and DNA methylation were represented by only up to two CpG sites at each of the genes, and the correlations were negative for *ATP5A, MFN2* and *NRF1* and positive for *NAPRT* and *Clp* protease (Figure 4F). In WAT, there were only six associations between the gene expression and CpG site methylation at the respective genes, three of which were annotated to *NAMPT* and one CpG to each of the other three genes *NMNAT3, ATP5A* and ubiquinol-cytochrome c reductase core protein 2 (Figure 4F). Although it is difficult to draw any firm conclusions on the effects of DNA methylation on gene expression, these results suggest that epigenetic control could be one of the mechanisms via which NR regulates the expression of genes participating in mitochondrial biogenesis and quality control, satellite cell stemness, NAD^+^ biosynthesis and OXPHOS.

### NR alters plasma metabolomic profile

To understand whether NR affects the global plasma metabolomic profile, we performed a nontargeted metabolomics analysis of fasting plasma samples from the twins of the BMI-discordant pairs. With regression analysis, we found 460 significantly altered metabolites upon NR. Amongst these, 72 metabolites were identified with a level 1 or 2 in the metabolomics standards initiative reporting standards (Table S7). Of the 72 identified metabolites, the most significantly increased metabolite was methylNAM (Figure 5), the NAM waste product, in line with the whole blood NAD metabolome (Figure 1F) and global DNA methylation results (Figure 4A-4B). In addition, the levels of tyrosine, the amino acid precursor of brain catecholamines, were significantly elevated upon NR treatment (Figure 5). Most of the decreased metabolites fell into the following categories: energy metabolism, amino acids, fatty acids, phospholipids and sphingomyelins. The content of metabolites central for the efficient mitochondrial energy production; carnitine, acylcarnitines and polyunsaturated fatty acids, decreased significantly (Figure 5) suggesting their lowered synthesis/excretion and/or increased uptake/utilization by the tissues. NR decreased the plasma levels of several amino acids including gluconeogenic amino acids (Figure 5) indicating also their diminished synthesis/excretion and/or increased uptake/utilization by the tissues such as the liver, which is the main tissue responsible for amino acid metabolism in the postprandial state (Rui, 2014). Given that the plasma levels of 3-carboxy-4-methyl-5-propyl-2-furanopropionic acid, a putative biomarker of fatty fish intake (Hanhineva et al., 2015), were significantly decreased (Figure 5), we cannot exclude the possibility that the lowered levels of polyunsaturated fatty acids partially reflected a decline in fish intake. The decrease in phosphatidylcholines and 1,5-diaminonaphthalene was more pronounced in the heavier cotwins of the BMI-discordant pairs (Table S8; on the 1 or 2 metabolite identification level). Overall, our results revealed that the main plasma metabolites modulated by the NR supplementation are related to mitochondrial energy metabolism, lipids and amino acids.

**Figure 5.**
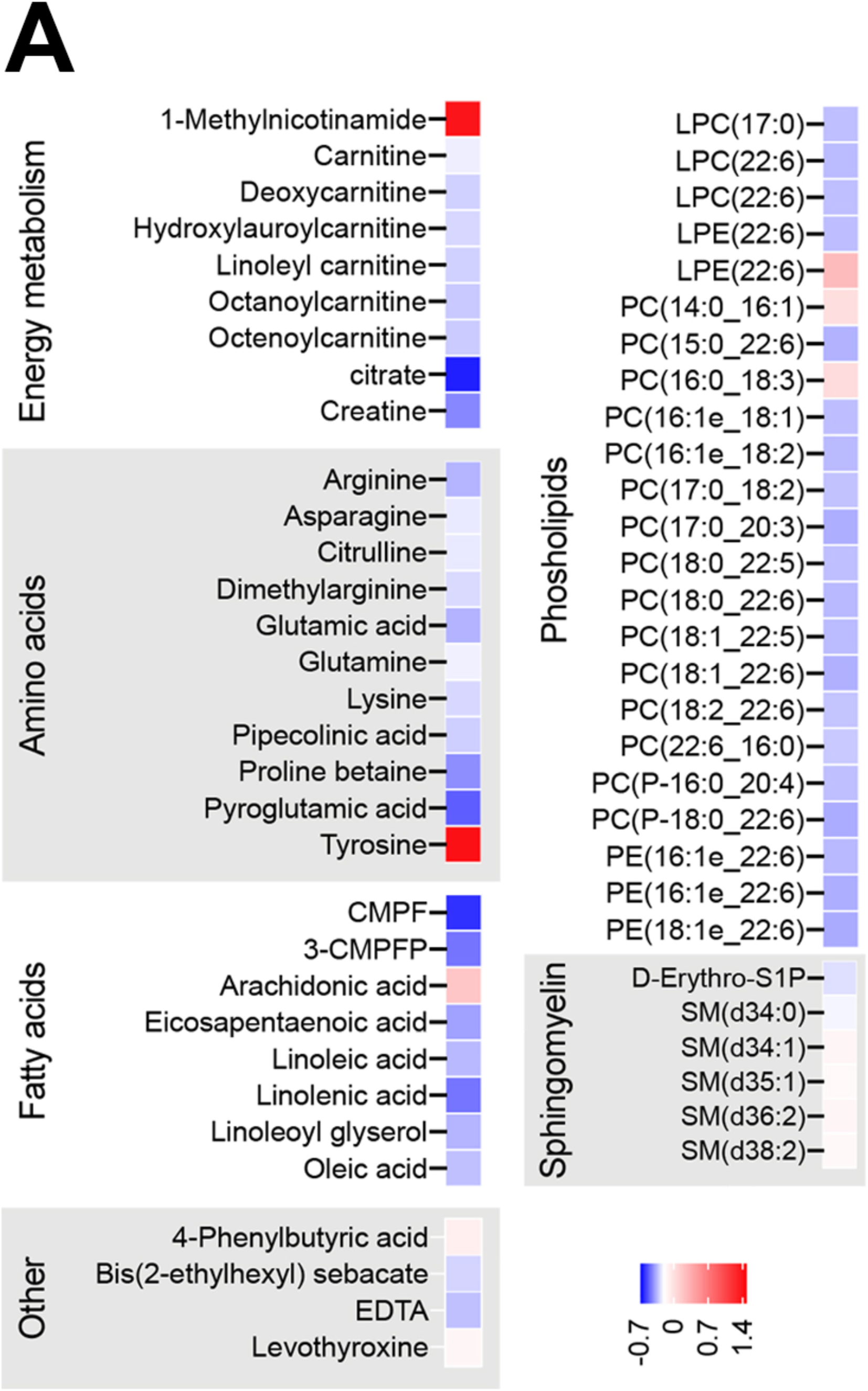
NR alters plasma metabolomics profile in the twins from the BMI-discordant pairs. **(A)** Heat maps showing significant plasma metabolite log2-fold changes post NR (n=14 twin pairs/28 individuals; nominal p< 0.05). Blue color indicates lower and red higher metabolite level upon NR. CMPF, 3-carboxy-4-methyl-5-propyl-2-furanopropionic acid; 3-CMPFP, 3-carboxy-4-methyl-5-pentyl-2-furanpropanoic acid; EDTA, Ethylenediaminetetraacetic acid; LPC, lysophosphatidylcholine; LPE, lysophosphatidyl-ethanolamine; PC, phosphatidylcholine; PE, phosphatidylethanolamine; D-Erythro-S1P, D-erythro-Sphingosine-1-phosphate; SM, sphingomyelin. Related statistical analyses are described under the Method section. See also Tables S7-S8.

### NR slightly modulated gut microbiota composition

To evaluate whether NR modified gut microbiota, 16S rRNA gene sequencing of fecal samples of the twins from the BMI-discordant pairs was performed. The alpha- or beta-diversity of the gut microbiota did not change during the study (Figures 6A-6B). In contrast, NR increased the abundance of the genus *Faecalibacterium* by ~2% but this increase did not pass the multiple testing correction via false discovery rate (FDR) (Figures 6C-6E). As *F. prausnitzii* is the only validated species of *Faecalibacterium genus,* we quantified the NR-induced changes in *F. prausnitzii* using quantitative PCR. Indeed, there was an increasing ~1.7 fold effect on *F. prausnitzii* upon NR (Figure 6F). The changes for any of the analyzed microbiota operational taxonomic units (OTUs) did not significantly differ between the heavier and the leaner cotwins of the BMI-discordant pairs (FDR p>0.05) (data not shown). Overall, our data suggest that NR increased the abundance of *F. prausnitzii,* which is one of the most important commensal bacteria of the human gut microbiota (Munukka et al., 2017; Sokol et al., 2008).

**Figure 6.**
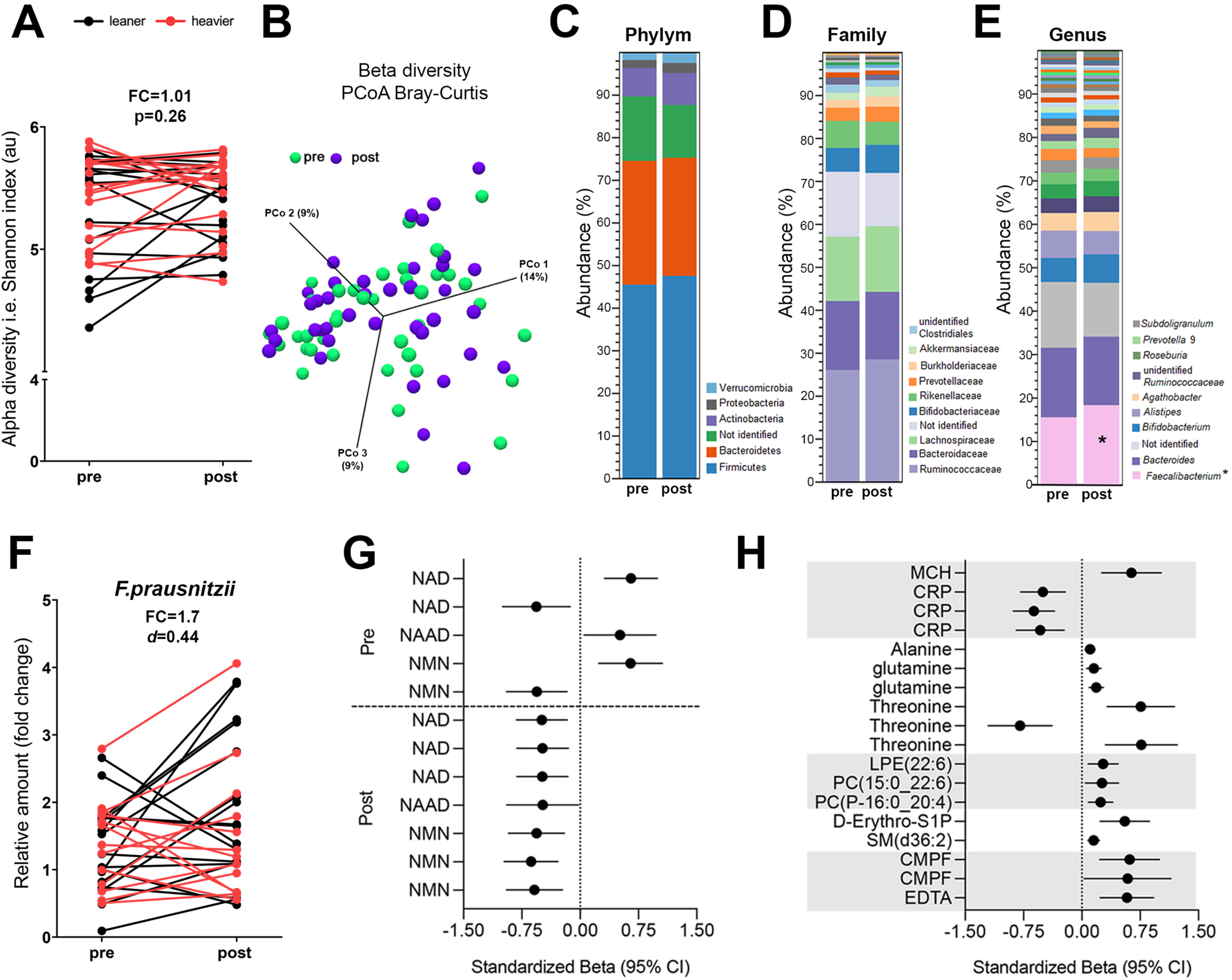
NR slightly improves gut microbiota composition in the twins from the BMI-discordant pairs. **(A)** Alpha-diversity (Shannon index *i.e*., species diversity) analysis of the gut microbiota pre versus post NR (n=11 twin pairs/22 individuals). **(B)** Principal Component analysis (PCoA) of the gut microbiota beta-diversity pre versus post NR (n=11 twin pairs/22 individuals). According to Bray Curtis distance and PERMANOVA analysis, NR treatment had no effect on the beta-diversity, *i.e.* inter-individual species diversity. **(C-E)** Average gut microbiota abundance at phylum (C), family (D) and genus (E) level pre versus post NR (n=11 twin pairs/22 individuals). In the legends on the right side of the bars, the phylum level shows all detected phyla in the samples, and at the family and genus level, the top 10 most abundant taxa are shown. * denotes a statistically significant change (nominal p< 0.05) upon NR. **(F)** *Q*uantification of *F. prausnitzii* amount in fecal DNA pre versus post NR using quantitative PCR (n=11 twin pairs/22 individuals). Cohen’s d was used to calculate changes in the *F. prausnitzii* amount. Cohen’s d between 0.5 and 0.8 suggests a medium effect size. Fold change (FC) indicates the mean of post NR value divided by the pre NR value. **(G)** Standardized coefficients (βs) showing associations (in standard deviations) between the baseline abundance of *Faecalibacterium* (pre) or the change of *Faecalibacterium* abundance with the change in NAD metabolites upon NR (post (n=11 twin pairs/22 individuals). Each row represents an association with *Faecalibacterium* operational taxonomic unit. Error bars denote 95% confidence intervals. Associations were considered significant when FDR< 0.05. **(H)** Standardized coefficients (βs) showing associations (in standard deviations) between the NR-induced changes in *Faecalibacterium* and blood clinical variables and plasma metabolites (n=11 twin pairs/22 individuals). Each row represents an association with *Faecalibacterium* operational taxonomic unit. Error bars denote 95% confidence intervals. Associations were considered significant when FDR< 0.05. Statistical analyses for all gut microbiota and association analyses are described in more detail under the Method section. See also Table S9.

### The abundance of Faecalibacterium associates with changes in blood metabolite concentrations

As the gut microbiota can influence the response of the host to vitamin B3 supplementation (Shats et al., 2020), we determined associations between OTUs and blood metabolites (Table S9) in the twins from the BMI-discordant pairs. At baseline, three out of five *Faecalibacterium* OTUs associated positively with the change in whole blood NAD^+^, NAAD and NMN upon NR indicating that the baseline abundance of *Faecalibacterium* may support the NR-induced elevation in blood NAD metabolome (Figure 6G). In addition, the NR supplementation-induced increase in *Faecalibacterium* abundance was negatively associated with the change in whole blood levels of NAD^+^, NAAD and NMN (Figure 6G) possibly reflecting competition for NR between *Faecalibacterium* and the host. The association analysis with blood clinical variables revealed that the *Faecalibacterium* increase was negatively associated with the change in high sensitivity CRP but positively with the change in mean corpuscular hemoglobin (Figure 6H). Lastly, the *Faecalibacterium* increase was associated positively with the change in plasma gluconeogenic amino acids (alanine, glutamine and threonine), phospholipids and sphingomyelins (Figure 6H). Altogether, our results indicate that *F. prausnitzii* may contribute to the regulation of NAD^+^, inflammation, and amino acid and lipid metabolism in humans.

### The effect of placebo- and NR-treatments in the twins from the BMI-concordant pairs

We lastly studied the impact of NR on whole blood NAD metabolites, body composition and clinical variables in comparison to placebo in four BMI-concordant twin pairs. We observed that NR treatment appeared to induce whole blood NAD^+^, NAAD and Me4py in comparison to placebo (Figures S2I-S2N) confirming the compliance to the NR treatment. Of the body composition parameters, body weight (~3 kg), fat percentage, liver fat percentage and adipocyte weight and volume increased (however, statistically non-significantly probably due to small sample size) in the placebo-treated cotwins while these variables remained stable in the NR-treated cotwins (Table S10). We did not observe any changes in the lifestyle factors, glucose and lipid metabolism and cardiovascular health-related variables in NR- or in placebo-treated twins from the BMI-concordant pairs (Table S10). In conclusion, based on our findings in the twins from the BMI-concordant pairs, NR’s effect on body composition and metabolic health did not markedly differ from the effects of placebo.

### Reported side-effects and safety parameters

NR supplementation was well-tolerated by the study participants. Reported side-effects included muscle pain, gastrointestinal irritation, sweating, nausea and headache but not cutaneous flushing typically caused by niacin (Ganji et al., 2014). The placebo-treated twins from the BMI-concordant pairs did not report any side effects. Safety parameters including for example the kidney function marker creatinine, liver enzymes and blood count remained unaltered in the blood samples of all study participants (Tables S10-S11). The only exception was that there was a statistically but not clinically significant reduction in hemoglobin in the twins from the BMI-discordant pairs upon NR (Table S11). However, this normocytic, normochromic condition (Table S11) was likely related to blood loss due to blood sampling. As a whole, these results showed that long-term NR supplementation did not markedly alter the measured safety parameters.

## Discussion

The boosting of NAD^+^ levels is currently under the focus of intensive research as a potential treatment option for metabolic diseases (Cantó et al., 2015). In our study, NR effectively increased the levels of whole blood NAD^+^, NAAD, NMN and Me4Py. This is in line with the previous NR clinical studies (Airhart et al., 2017; Dollerup et al., 2018; Elhassan et al., 2019; Martens et al., 2018; Remie et al., 2020; Trammell et al., 2016a). In comparison with niacin, NR with the dose of 1g/day was less potent in elevating whole blood NAD^+^ levels as NR induced a 2-fold increase after 5 months while niacin with the same dose induced a 6-fold increase after 4 months (Pirinen et al., 2020). However, the levels of NR and niacin were not molar equivalents in these studies; the molar concentration was higher for niacin. So far, short-term clinical studies have not reported the positive effects of NR on tissue NAD^+^ biosynthesis (Airhart et al., 2017; Dollerup et al., 2018; Elhassan et al., 2019; Martens et al., 2018; Remie et al., 2020; Trammell et al., 2016a). We here provide the first evidence that long-term NR supplementation increases muscle and WAT NAD^+^ biosynthesis in humans regardless of BMI. This NAD^+^ biosynthesis boosting effect occurred even in the heavier cotwins from the BMI-discordant pairs, which had downregulation of NAD^+^ biosynthetic genes, *NMNAT3 and NRK1,* especially in WAT, at baseline. Given that the upregulation of both salvage pathway enzymes *NRK1* and *NAMPT* was observed upon NR, especially in muscle, NAD^+^ generated in tissues was likely originating both from NR and NAM, the degradation product of NR. This agrees with mouse studies showing that most of the orally consumed NR is metabolized to NAM and a small proportion of NR is directly utilized for NAD^+^ biosynthesis via NRKs (Liu et al., 2018).

Currently, there is plenty of evidence for the efficacy of NR on muscle mitochondrial biogenesis in mice (Canto et al., 2012; Cerutti et al., 2014; Frederick et al., 2016; Yu et al., 2021; Zhang et al., 2016) but not in humans (Dollerup et al., 2020; Remie et al., 2020). In our study, the NR-induced improvement in NAD^+^ metabolism led to an increase in muscle mitochondrial number and mtDNA amount after 5 months. In line with the previous mouse studies (Canto et al., 2012; Wang et al., 2018), the mechanism was likely mediated via *ESRRA/TFAM/MFN2*. Collectively, our evidence emphasizes that a supplementation time longer than 3 months is required to detect the impact of NR on muscle mitochondrial biogenesis in healthy individuals with overweight and obesity. This notion is supported by our previous finding showing that niacin with the dose of 1 g/day significantly increased muscle mitochondrial mass in healthy normal-weight controls after 4 months (Pirinen et al., 2020). Overall, our current clinical trial is a proof-of-principle of NR’s effects on muscle mitochondrial biogenesis in humans.

NR is known to exert beneficial effects on muscle satellite cells in mice (Zhang et al., 2016). Here, we show that NR reduced the number of PAX7^+^ satellite cells likely by increasing differentiation and fusion of satellite cells to the existing myofibers. Given that we did not observe the increase in muscle mass, our data reveal that satellite cell fusion can be uncoupled from muscle hypertrophy under specific conditions in humans. It has been shown previously that during homeostasis, reduced Notch activity in satellite cells results in premature differentiation and fusion to myofibers bypassing self-renewal (Mourikis et al., 2012). It will be interesting to probe if NR modulates Notch signaling in satellite cells. Our myoblast results align with a recently published study demonstrating an increased differentiation capacity of NR-supplemented human muscle precursor cells (Seldeen et al., 2021). One molecular mechanism mediating the NR’s effect on myogenic differentiation could be the NR’s degradation product, NAM, which is shown to increase myogenesis in mouse stem cells (Zhou et al., 2015). Whether the increased differentiation of satellite cells provides a functional benefit or disadvantage upon exercise and muscle damage needs to be addressed in future studies.

In mice, NR with the dose of 400-500 mg/kg/day has been shown to counteract the negative metabolic consequences of a high-fat diet (Canto et al., 2012), but a high dose of NR (~1000 mg/kg/day) to cause WAT dysfunction and impaired glucose homeostasis (Shi et al., 2019). So far, positive outcomes on body composition from the clinical trials are largely lacking (Airhart et al., 2017; Dollerup et al., 2018; Elhassan et al., 2019; Martens et al., 2018; Remie et al., 2020; Trammell et al., 2016a). During our study, adiposity increased in the twins from the BMI-discordant pairs although their self-reported lifestyle factors remained the same. The underlying mechanism may be linked to morphological and molecular changes in WAT. However, we did not detect changes in adipocyte number or size or WAT *PPAR*γ dysregulation that has been reported in mice upon high doses of NR (Shi et al., 2019). As the body weight and fat mass tended to increase also in the placebo-treated twins from the BMI-concordant pairs, the weight gain may be related to a normally occurring increase in adiposity over time rather than to NR *per se* (Chen et al., 2019). Interestingly, in our previous study, the long-term niacin supplementation resulted in an opposite outcome related to body composition compared with the effects of NR in the current study. Niacin with the dose of 1 g/day reduced whole-body fat percentage in normal-weight study participants after 4 months (Pirinen et al., 2020). Similarly, as for adiposity, no significant effect on insulin sensitivity has been found in the published clinical trials with NR (Dollerup et al., 2018; Martens et al., 2018; Remie et al., 2020). Here we report elevated fasting glucose and insulin levels and impaired insulin sensitivity in the OGTT in twins from the BMI-discordant pairs after 5-month NR supplementation. As niacin is well known to raise fasting glucose levels and impair insulin sensitivity by increasing hepatic gluconeogenesis and by decreasing insulin signalling (Guyton and Bays, 2007), our findings raise the question whether niacin and NR exhibit a similar mechanism of action on glucose metabolism in humans. Overall, based on the current clinical evidence, larger and longer placebo-controlled studies are required to clarify the role of NR as a modifier of adiposity and glucose metabolism in humans. Given that most clinical studies with NR, like ours, have been performed in metabolically healthy individuals (Airhart et al., 2017; Elhassan et al., 2019; Martens et al., 2018; Remie et al., 2020; Trammell et al., 2016a), human studies in patient populations with metabolic diseases are warranted in the future.

Epigenetic mechanisms such as DNA methylation have been shown to regulate gene expression upon NR in mice (Serrano et al., 2020). Typically, DNA methylation in gene promoter regions is associated with transcriptional repression. However, the relationship between DNA methylation and gene expression at a given location is not straightforward. Here, we demonstrate that NR supplementation led to a muscle-specific decline in global DNA methylation likely via reduced methyl pool size due to increased elimination of NAM via methylation. Nevertheless, both hyper- and hypomethylation of CpG sites were detected in muscle and WAT although most of the analyzed sites were hypomethylated upon NR. Notably, methylation changes at the CpG sites were associated with the altered tissue expression for several analyzed genes such as *PGC-1α*, *PAX7* and *NAMPT*. Therefore, our study is the first demonstration that the epigenetic mechanisms may be involved in the reprogramming of tissue NAD^+^ and mitochondrial metabolism and muscle stem cell identity upon NR in humans. The mechanism via which NR diminishes DNA methylation could be related to the inhibition of DNA methyltransferases through direct or indirect mechanisms as suggested previously (Serrano et al., 2020).

The disruption of gut microbiota homeostasis *i.e.* dysbiosis is a typical feature of metabolic and muscle diseases (Fan and Pedersen, 2021). In line with the previous mouse studies (Jiang et al., 2020; Yu et al., 2021), NR ingestion slightly improved microbiota composition in our study participants. Similarly, niacin has been reported to beneficially affect the gut microbiota composition in humans (Fangmann et al., 2018). We here show that NR supplementation increased the proportion of the only identified species of the genus *Faecalibacterium,* namely *F. prausnitzii,* which is a gut bacterium that promotes metabolic health and anti-inflammatory responses (Munukka et al., 2017; Sokol et al., 2008). In agreement with the latter notion, the *Faecalibacterium* abundance was negatively associated with high sensitivity CRP. Intriguingly, our correlation analyses between the *Faecalibacterium* abundance and whole blood NAD metabolites imply that *F. prausnitzii* may compete for the NAD^+^ precursors with the host for its own NAD^+^ biosynthesis and promote the *in vivo* efficiency of NR in humans. Supporting these conclusions, it is known that *F. prausnitzii* can metabolize NR and NMN (Heinken et al., 2014) and that gut microbiota are crucial for the NAD^+^ boosting effect of NR in mice (Shats et al., 2020). Taken together, our findings suggest that NR changes gut microbiota composition in a manner that may improve health and that the NAD^+^ metabolism could be partly regulated through the interaction between the host and *Faecalibacterium* in humans.

In conclusion, NR supplementation is a potential treatment option to be tested in individuals with decreased muscle mitochondrial biogenesis and dysbiosis. As possible adverse effects, we report a declining muscle satellite cell number and a possibility of impaired glucose metabolism. As patients with chronic muscle disease typically exhibit satellite cell dysfunction, muscle performance and regeneration are important endpoints to be monitored in future NR clinical trials. Overall, our data underscore the role of NR as a potent modifier of systemic NAD^+^ levels, muscle mitochondrial biogenesis, satellite cell function, DNA methylation and microbiota in humans. Notably, BMI does not seem to be the important factor affecting the response to NR in humans.

## Limitations of the study

The study participants volunteered to two muscle and WAT biopsy collections yielding material sufficient for histology, electron microscopy, RNA and DNA isolation but not for mitochondrial respirometry and protein level analyses. The electron microscopic evaluation of tissues was limited to muscle due to improper fixation of WAT samples. The open study setting and the lack of a statistically powered placebo group may compromise our results related to body composition and metabolic health, and these endpoints need to be followed up in larger placebo-controlled trials. However, our rigorous molecular analyses unequivocally indicate that long-term NR supplementation impacts muscle mitochondrial biogenesis, stem cell responses, as well as DNA methylation and microbiota composition.

## STAR★Methods

Detailed methods are provided in the online version of this paper and include the following:

- KEY RESOURCES TABLE
- RESOURCE AVAILABLILTY

- LEAD CONTACT The Lead Contact is Eija Pirinen
- MATERIALS AVAILABILITY This study did not generate new unique reagents. Requests for resources and reagents should be directed to the Lead Contact.
- DATA AND CODE AVAILABILITY All data are part of the ‘Twin Study’ and are deposited with the Biobank of the Finnish Institute for Health and Welfare (https://thl.fi/en/web/thl-biobank/for-researchers/sample-collections/twin-study). For details on accessing the data, see https://thl.fi/en/web/thl-biobank/for-researchers/application-process. All bona fide researchers can apply for the data.
- EXPERIMENTAL MODEL AND SUBJECT DETAILS

- Study participants
- METHODS DETAILS

- Subject recruitments and study design
- Body composition and energy expenditure
- Blood laboratory examinations
- Adipose tissue sampling
- Muscle sampling and histology
- Myoblast cell cultures
- Targeted quantitative NAD metabolome analysis
- mtDNA analyses and quantification
- Gene expression analyses
- DNA methylation analysis
- Plasma metabolite isolation and non-targeted metabolite profiling
- Plasma metabolite identification and differential global metabolite profile analysis
- Gut microbiota sequencing and composition analyses
- Association analyses
- QUANTIFICATION AND STATISTICAL ANALYSIS

- Statistical Analysis
- ADDITIONAL RESOURCES

## Data Availability

All data are part of the Twin Study and are deposited with the Biobank of the Finnish Institute for Health and Welfare (https://thl.fi/en/web/thl-biobank/for-researchers/sample-collections/twin-study). For details on accessing the data, see https://thl.fi/en/web/thl-biobank/for-researchers/application-process. All bona fide researchers can apply for the data.

## Acknowledgements

We thank the study participants for their valuable contribution to this research. We acknowledge Mia Urjansson, Katja Sohlo, Tarja Hallaranta, Antti Muranen, Ilse Paetau, Helinä Perttunen-Nio, Elisa Silvennoinen, Anna Sandelin, Saila Saarinen and Minna Tuominen for their technical expertise. We also thank Mervi Lindman and Ilya Belevich (the Electron Microscopy Unit of the Institute of Biotechnology, University of Helsinki) for the TEM sample preparation and help in preprocessing and analyzing the TEM images, respectively. We wish to acknowledge the core facility services Genome Biology Unit, HiLIFE and Faculty of Medicine, University of Helsinki, and Biocenter Finland and Light Microscopy Unit, Institute of Biotechnology, University of Helsinki. We also would like to thank Ryan Dellinger and the Chromadex CERP Science Team.

The study was supported by Centre of Excellence funding from the Academy of Finland (#272376 KHP and #336823 JK), as well as the Academy of Finland grants (286359 and 314455 EP, 335443, 314383, 272376 and 266286 KHP, 297908 and 328685 MO), the Sigrid Jusélius Foundation, the Finnish Medical Foundation, the Medical Society of Finland, the Finnish Diabetes Research Foundation, the Novo Nordisk Foundation (NNF20OC0060547, NNF17OC0027232, NNF10OC1013354), the Gyllenberg Foundation, the Orion Research Foundation, the Biomedicum Helsinki Foundation, Government Research Funds (Helsinki University Hospital). This research was also supported by the Academy of Finland Profi6 336449 funding.

## Author Contributions

E.P. and K.H.P. designed the study and supervised the project. E.P., H.L., M.K. and A.He. wrote the manuscript. H.L., M.K., A.He., S.P. and B.W.v.d.K. prepared figures and tables. M.K. S.S., N.P., S.G., and M.S. conducted wet-lab experiments. H.L., M.M., B.W.V., A.He. and S.P. performed data analyses. K.H.P. recruited the study subjects and studied them together with H.L., S.H., T.S., J.Ka. provided the twin cohort. M-RT contributed to the analysis of blood lipid profiles and P.K. to satellite cell studies. M.O. was responsible for DNA methylation analysis and M.T. and S.P. for gut microbiota analysis. N.L., A.Ha., J.Ku., and P-H.G. contributed to the body composition analyses. M.L. and J.T. performed plasma global metabolomics analysis, and C.B. and M.S. NAD^+^ metabolome analysis. All authors revised and approved the manuscript. E.P., H.L., M.K., A.He, B.W.v.d.K., S.P., M.M., M.O. and K.H.P. interpreted the data.

## Experimental Model and Subject details

### Study participants

This study was approved by the Ethics Committee of the Helsinki University Central Hospital (protocol number 270/13/01/2008) and the study was conducted according to the principles of the Declaration of Helsinki. The study was registered at clinicaltrials.gov with the entry NCT03951285. Written informed consent was obtained from all subjects. Twenty-six MZ twin pairs belonging to the Finnish Twin Cohort study were screened for eligibility for the current study. Twenty-two MZ twin pairs with the cotwins having a BMI difference of at least 2.5 kg/m^2^ were considered as BMI-discordant. Two such twin pairs did not meet the inclusion criteria, as shown in Figure S1. Another four twin pairs were considered as BMI-concordant (within-pair difference in BMI <2.5 kg/m^2^). The twins were 29-65 years old, with a mean age of 40. Fourteen of the MZ twin pairs were female and twelve were male pairs. The study participants had the following medications: hypertension (n=2; angiotensin II receptor blocker, calcium channel blocker, diuretic), asthma or allergy (n=7; an inhaled corticosteroid, beta-2-agonist, antihistamine), type 2 diabetes (n=1; metformin, insulin), rheumatoid arthritis (n=2; hydroxychloroquine, methotrexate), psychiatric (n=3; selective serotonin intake inhibitor, valproate, a tricyclic antidepressant, neuroleptic), oral contraceptives (n=2). The number of smokers is shown in Table S1.

Inclusion criteria were as follows: (i) age >18 years; (ii) BMI >18.5 kg/m^2^ in both members of the twin pair; (iii) agreement to maintain current level of physical activity throughout the study; (iv) agreement to avoid vitamin supplementation or nutritional products with vitamin B3 14 days prior to the enrolment and during the study.

The exclusion criteria were: (i) unstable medical conditions as determined by the investigator; (ii) clinically significant abnormal lab results at screening (e.g. aspartate aminotransferase and/or ALT > 2 × upper limit of normal, and/or bilirubin > 2 × upper limit of normal); (iii) subjects who would have had a planned surgery during the course of the trial; (iv) History of or a current diagnosis of any cancer (except for successfully treated basal cell carcinoma diagnosed less than 5 years prior to screening). Subjects with cancer in full remission more than 5 years after diagnosis were acceptable; (v) history of blood/bleeding disorders; (vi) immunocompromised individuals such as subjects that had undergone organ transplantation or subjects diagnosed with human immunodeficiency virus; (vii) hepatitis; (viii) blood donation in the previous 2 months.

## Method Details

### Subject recruitment and study design

BMI-discordant and BMI-concordant MZ twin pairs were screened from two large population-based cohorts of Finnish twins (FinnTwin12, Finntwin16, n=5000, 1200 of which MZ), established by Professor J.K. (Kaprio, 2013) and recruited by K.H.P. The procedures for the selection of study participants are described in Figure S1. One-to-one phone conversations were conducted at least once a month to follow up compliance to the study and potential symptoms or side effects. All participants were instructed to continue their normal routine and not make any changes to their physical activity and diet.

The examination protocol of this non-randomized, open-label study is described in Figure 1A. All twins from the BMI-discordant pairs received NR supplementation. For BMI-concordant twin pairs, one cotwin received NR and the other cotwin received placebo. The cotwins were allocated to the two groups randomly and in a blinded fashion. The NR dose, 1 g/day, was selected based on a previous study showing that 1 g/day niacin (another form of vitamin B3) supplementation had a favorable effect on systemic NAD^+^ levels, body composition and mitochondrial biogenesis (Pirinen et al., 2020). The weekly dose was initially 250 mg/week. The daily NR dose was gradually escalated by 250 mg/week so that the full dose of 1 g/day was reached in one month. The supplementation was continued for four months with the full dose. At the end of the study, the dose was slowly decreased to the initial level at a 250 mg/week rate. The collection of samples such as blood, WAT and muscle biopsies, saliva and feces was performed at the baseline and at the end of the study (*i.e.* 5 months). Four BMI-discordant twin pairs discontinued the intervention as described in Figure S1. Sixteen BMI-discordant and four BMI-concordant twin pairs were included in the further analyses, as shown in Figure S1. Safety laboratory tests included blood count, iron levels, and liver and kidney function tests. The analysis of body composition, clinical variables and whole blood NAD^+^ metabolites was performed for all study participants. The molecular measurements including gene expression, imaging and omics analyses were conducted only for the samples from the BMI-discordant twin pairs.

### Body Composition, Energy Intake and Expenditure

Weight, height, waist-circumference, whole-body fat (assessed by using dual-energy x-ray absorptiometric scans), abdominal subcutaneous and visceral fat (by magnetic resonance imaging, MRI), and liver fat (by magnetic resonance spectroscopy, MRS) were measured as described previously in all study participants (Graner et al., 2012). In short, MRI/MRS for amounts of subcutaneous adipose tissue, visceral adipose tissue, and liver fat were performed on a clinical 1.5-T imager (Avanto, Siemens, Erlangen, Germany). To allow measurement of abdominal fat distribution, a stack of abdominal T1-weighted MR images (16 slices, slice thickness 10 mm, TR of 91 ms, TE of 5.2 ms and flip angle of 80°) were obtained from 8 cm above to 8 cm below the L4/5 lumbar intervertebral disks using frequency selective fat excitation. The amount of visceral and subcutaneous adipose tissue was determined from each slice using segmentation software SliceOmatic 5.0 (TomoVision, Quebec, Canada) utilizing region growing routine.

In turn, a point resolved spectroscopy sequence was used for volume selection in hepatic MRS. A 25×25×25 mm^3^ voxel was placed in the middle of the right liver lobe and liver spectra with TE of 30 ms and four averages were collected. Signal acquisition was triggered to end exhalation using a navigator belt to eliminate motion artifacts due to respiratory motion so that TR was kept >4000 ms. Liver spectra were analyzed with jMRUI 6.0 software (Stefan et al., 2009) and intensities of methylene and water resonances were determined using the AMARES algorithm (Vanhamme et al., 1997). Signal intensities were corrected for relaxation effects and liver fat was determined as an intensity ratio of methylene/(methylene+water). Ratios were further converted to mass fractions as described previously by (Kotronen et al., 2009).

Muscle mass and energy expenditure were measured via bioelectrical impedance analysis using the Tanita MC-980 device. Physical activity was measured using the Baecke questionnaire (Baecke et al., 1982) and dietary intake was assessed from 3-day food records and analyzed with the Diet32 program (Aivo Finland Oy), based on a national Finnish database for food composition (Fineli, www.fineli.fi).

### Blood Laboratory Examinations

Routine blood tests were performed for all study participants. Blood samples were collected after overnight fasting, and whole blood, separated plasma and serum samples were frozen at −80C. Blood count, ALT, aspartate aminotransferase, HbA1c, glucose, insulin, C-peptide, lactate, total cholesterol, HDL, LDL, triglycerides, homocysteine and high-sensitivity CRP were analyzed using standardized methods at the HUSLAB laboratories.

A 75-g oral glucose tolerance test with 4-time points (0, 30, 60, and 120 min) was performed after a 12-hour overnight fast, followed by measurements of plasma glucose with spectrophotometric hexokinase and glucose-6-phosphate dehydrogenase assay (Roche Diagnostics) and serum insulin with time-resolved immunofluorometric assay (PerkinElmer). Plasma HDL was measured as cholesterol after precipitation of other lipoproteins with heparin–Mn2^+^ (Bachorik and Albers, 1986). Free fatty acids were determined using NEFA kit (Wako Chemicals), apolipoprotein B levels were determined with Apolipoprotein B Konelab Kit (Thermofisher Scientific), while plasma adiponectin levels were measured using ELISA kit (R&D Systems).

### Adipose Tissue Sampling

Adipose tissue biopsies were collected from all study participants at baseline and at the end of the study (Figure 1A). The subcutaneous WAT biopsies were taken from superficial abdominal adipose tissue near the umbilicus using a needle biopsy under local anesthesia (lidocaine). WAT samples for RNA, DNA and metabolite analyses were snap frozen and stored in liquid nitrogen or −80°C until used.

Samples used for determination of the adipocyte size and number were, in turn, immediately processed by collagen digestion and separation of adipocytes by centrifugation as described previously by (Heinonen et al., 2014). In brief, the subcutaneous adipose tissue was minced and incubated for 1 h at 37 °C with continuous shaking in 10 ml of adipocyte medium (DMEM/F-12 (1:1) (Invitrogen) with 16 μmol/l biotin, 18 μmol/l panthotenate, 100 μmol/l ascorbate and antibiotic-antimycotic (Invitrogen)), supplemented with 2% bovine serum albumin (Sigma) and with 2 mg/ml collagenase A (Roche Diagnostics). Digestion was terminated by adding adipocyte medium supplemented with 10% newborn calf serum (Sigma) and centrifuging for 10 min at 600 × g. After washing the adipocytes with adipocyte medium, images of the adipocytes were taken with a light microscope (Zeiss, Axioplan2) using 50X magnification. Adipocyte diameters were automatically measured from the images using a custom algorithm for ImageJ (ImageJ 1.42q/ Java 1. 6.0 10 32-bit; https://github.com/birgittavdkolk/vanderkolk_etal_2021) (van der Kolk et al., 2021) which preprocessed the image to enhance the borders of the adipocytes and then used a circle-detection algorithm to identify the cells. The algorithm was tuned to identify the adipocytes taken using the standardized microscope settings and validated against 2000 manually measured diameters from 20 pictures (r = 0.85, p < 0.001). Mean adipocyte volume was calculated for each individual using the following formula:

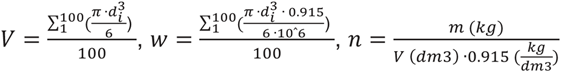

*V* = cell volume (μm^3^), *d* = cell diameter (μm), w=cell weight (μg), n = total adipocyte number, where m = total body-fat mass (kg) and V (m^3^)*rho (density, fat) = mean weight of a single adipocyte. Density of fat cell triglycerides = 0.915 g/ml. Adipocytes were assumed to be spheres.

### Muscle Sampling, TEM, and Satellite Cell Histology and Quantification

Muscle tissue biopsies were collected at baseline and at the end of the study from all study participants (Figure 1A). A Bergström needle biopsy from the vastus lateralis muscle was taken under local anesthesia in sterile conditions by making an incision through the skin and underlying fascia. The muscle samples for RNA, DNA and metabolite analyses were snap frozen and stored in liquid nitrogen or at −80°C.

Samples for TEM analysis were fixed in 2.5% glutaraldehyde. For plastic embedding, they were then treated with 1% osmium tetroxide dehydrated in ethanol and embedded in epoxy resin. One mm section was stained with methylene blue (0.5% w/v) and boric acid (1% w/v). These methylene blue sections were used to mark the interesting areas for further TEM analyses by examining the sections with a light microscope. Thereafter, ultrathin (60– 90 nm) sections were cut on grids and stained with uranyl acetate and lead citrate by the Viikki Electron Microscopy Unit of the Institute of Biotechnology (EMBI, Biocenter Finland) and viewed with JEM-1400 transmission electron microscope (Jeol). From each study subject, 15 TEM images (both pre and post NR) were taken from the intermyofibrillar area of type I muscle fibers, determined by mitochondria content, using a 2000X magnification. Total mitochondria number per 10 µm^2^ relative to the muscle fiber area, mitochondrial surface area per muscle fiber area (%), dimension values (average diameter (µm) and perimeter (µm)), and total muscle fiber area (µm^2^) were determined by painting each mitochondrion in Microscopy Image Browser (MIB) (Belevich et al., 2016). Further, these values were used to calculate the total mitochondria number per 10 µm^2^ relative to the muscle fiber area, mitochondrial surface area per muscle fiber area (%, mitochondria density), and major shape descriptors, branching (form factor) and length-to-width ratio (aspect ratio).

Samples for muscle histological stainings were, in turn, snap frozen in liquid nitrogen-cooled isopentane. Cryosections from these samples were used to measure PAX7 immunostaining in muscle tissues: 9 µm thick frozen sections were post-fixed in 4% paraformaldehyde in DPBS for 10 min at RT and washed three times for 5 min in DPBS. Slides were then incubated in primary antibody (mouse monoclonal Pax7, Developmental Studies Hybridoma Bank (DSHB, Pax7 was deposited to DSHB by Kawakami A), 1:100 dilution in blocking solution (15% normal goat serum, 1.5% BSA, 0.5% Triton X-100 in 1X DPBS) overnight at 4°C. Thereafter, slides were washed three times for 10 min in DPBS and incubated in secondary antibody (Cy3 goat anti-mouse IgG1, 1:500 dilution, Jackson ImmunoResearch) and 1 μg/ml Hoechst for 1h at RT in order to visualize nuclei. Slides were washed as described above and mounted. Images were generated using Pannoramic 250 FLASH II digital slide scanner (3DHistec, Thermofisher Scientific) and Leica SP8 STED confocal microscope (Leica). For PAX7+ cell quantification, muscle sections were scored from a minimum of 2-3 sections/sample/condition and normalized to cross-sectional area of the muscle section scored. All quantifications were performed using Case Viewer 2.0 software (3DHistech, Thermofisher Scientific) and images were blinded for quantification. Details of the antibodies used are listed in the Key resources table.

### Myoblast Cell Cultures

Muscle samples for primary muscle cell cultures, *i.e*. myoblast cultures were processed immediately after excision. In short, samples were washed with 5 ml isolation medium (0.05% trypsin-ethylenediamine tetraacetic acid (EDTA) (SAFC Biosciences), 1.8 g/l glucose (Sigma), 7.15 g/l HEPES (Sigma), 7.6 g/l NaCl (Sigma), 0.224 g/l KCl (Sigma), 1.2 mg/l phenol red (Sigma)). Thereafter, 5 ml isolation medium was added, and tissue was minced. Another 10 ml of isolation medium was added and the sample was transferred to a 20 ml beaker with a magnetic stirrer and placed in a 37°C incubator for 20 min. The supernatant containing muscle cells was transferred into a 50 ml centrifuge tube containing 15 ml of myoblast growth medium (Ham’s F10 with Glutamax (Gibco) supplemented with 15% fetal bovine serum (FBS, Thermofisher Scientific), 500 mg/l fetuin (Sigma), 200 mg/l insulin (Sigma), 0.05% bovine serum albumin (BSA, Sigma), 0.39 mg/l dexamethasone (DMSO) (Sigma), 50 mg/l gentamicin (Sigma), 2 mg/l human epidermal growth factor (Thermofisher Scientific)). This process was repeated a further two times to break down any remaining tissue. Finally, the supernatant containing the isolated muscle cells was centrifuged for 10 min at 100 × g to collect the cells and the resulting cell pellet was resuspended in 5 ml of myoblast growth medium. Myoblasts were first seeded on 25 cm^2^ flasks (Corning). After reaching 70% confluence, they were subcultured in 150 cm^2^ flasks (Corning) and finally on six-well plates (Corning) at a density of 100000 cells per well. When confluent, they were collected for RNA isolation and stored at −80°C.

### Targeted quantitative NAD metabolome analysis

The measurement of whole blood NAD^+^ metabolome was carried out from all study participants after dual extractions as recently described (Pirinen et al., 2020). For analysis of Me4PY (group A analyte), samples were spiked with 400 pmol of [d3]-Me4PY (internal standard A). For analysis of NAD+, NADP, NMN, NAR, NAAD and ADPR, samples were dosed with 13C-yeast extract (internal standard B) as described (Elhassan et al., 2019; Trammell and Brenner, 2013).

In short, to quantify Me4PY (group A analyte), 75 μl of whole blood was added to 20 μL group A internal standard and 500 μl of 3:14%trichloroacetic acid (TCA):acetonitrile was added. The mixture was allowed to sit on ice for 20 min, after which samples were sonicated twice for 20 seconds and centrifuged at 4°C for 13 minutes at 16 100 × g. Next, the supernatant was removed and dried under vacuum overnight at room temperature. The samples were reconstituted in 2% acetonitrile/water immediately prior to the analytical run. To quantify NAD^+^, NADP, NMN, NAR, NAAD and ADPR (group B analytes), 75 μl of whole blood was added to 20 μl group B internal standard prepared in water and mixed with 500 μl of 3:1 4% (TCA):acetonitrile with vortexing. After resting on ice, the samples were centrifuged as described above. Next supernatant was removed and dried under vacuum overnight, and reconstituted in 2% acetonitrile/water. After reconstitution, samples were transferred to Waters polypropylene plastic total recovery vials and stored in a Waters Acquity H class autosampler maintained at 8°C until injection. In all cases, 8 μl of extract was loaded onto the column. For one set of samples, group B analytes plus Me4PY were extracted as for the group B analytes. 100 μl of blood was used and 200 pmol d3-Me4PY was added to the internal standard mix. All analytes in this group except NAAD had a corresponding stable labeled internal standard. Labeled NAD^+^ was used as the internal standard for NAAD.

Separation and quantitation of analytes were performed with a Waters Acquity LC interfaced with a Waters TQD mass spectrometer operated in positive ion multiple reaction monitoring mode as described (Trammell and Brenner, 2013) with minor modifications. MRM transitions monitored were: NAD^+^ 664.1>136.1; 13C10-NAD^+^ 674.1>136.1; NADP 744.1>136.1; 13C10-NADP 754.1>136.1; NAAD 665.1>136.1; NMN 335>123: 13C5-NMN 340>123; NAR 256>124; 13C5-NAR 261>124; ADPR 560>136.1; 13C10-ADPR 570>136.1; Me-4-py 153>136; d3-Me-4-py 156>139. Separate Hypercarb columns (Thermo Scientific) were used for the separations. The conditions for group A were solvent A 10 mM NH4OAc with 0.1% formic acid, solvent B acetonitrile with 0.1% formic acid, solvent D methanol, flow 0.30 mL/min; gradient initial 98% A, 2% B; 2.25 min, 98% A 2%B; 11 min, 74.8% A 17.9% B 7.3% D; 11.1 min,10% A 90% B; 14.3 min 10% A 90% B; 14.4 min, 98.2% A 2%B; end 18.5 min. Conditions for Group B were solvent A 7.5mM NH4OAc with 0.05% NH4OH, solvent B acetonitrile with 0.05% NH4OH; flow 0.353 ml/min; gradient initial 97% A 3% B; 1.8 min, 97% A 3% B; 10 min 65.5% A 34.5% B; 11 min 10% A 90% B; 13.2 min, 10% A 90% B; 13.3 min, 97% A 3% B; end 19 min.

### mtDNA Analyses and Quantification

Total DNA, including mtDNA, was extracted from the snap-frozen muscle and WAT samples (15 mg and 150 mg, respectively) using standard phenol–chloroform extraction and ethanol precipitation method, and stored at +4°C until use. mtDNA content was determined by analyzing the ratio of mtDNA amount relative to genomic DNA using SYBRGreen-based quantitative PCR (qPCR). To detect genomic DNA, primers for nuclear amyloid β precursor protein (*APP*), beta-2-microglobulin (*B2M*) and hemoglobin subunit beta (*HBB*) were used. In turn, to measure mtDNA, mitochondrial 16S rRNA (*16S*), cytochrome B (*CYTB*) and 12S rRNA (*DLOOP*) primers were chosen. The PCR primers are provided in the Key resources table.

qPCR was performed using 1X SYBRGreen MasterMix (Thermofisher Scientific), 10 µM of primers and 2 ng of DNA template in 384 well plate format using CFX384 Touch™ Real-Time PCR Detection Systems (Bio-Rad Laboratories). Thermal cycling as triplicates was run using initial denaturation of 3 min at 95°C, 39 cycles of 10 s at 95°C and 30 s at 62°C, final extension of 10 s at 95°C and melting curve analysis step from 65°C to 95°C (raised gradually in 0.5°C increments). The mtDNA amount was presented as a relative level of mtDNA genome per nuclear genome. Data analysis was performed using the ΔΔCt method with the qBASE+ 3.2 software (Biogazelle).’

### Gene expression

Total RNA from myoblasts was isolated using TRIzol reagent (Invitrogen) according to the manufacturer’s instructions. In turn, total RNA from muscle tissue and WAT was isolated using AllPrep DNA/RNA/miRNA Universal Kit (Qiagen) according to the manufacturer’s instructions with a few minor modifications. In short, muscle tissue (30 mg in 600 µl Buffer RLT Plus) was lysed using Tissuelyser II (Qiagen) at 25 Hz for 2 min and the lysate was transferred into the AllPrep DNA Mini spin column. WAT (250 mg in 1 ml Buffer RLT Plus), in turn, was lysed using Tissuelyser II (Qiagen) at 25 Hz for 2 × 2 min. As the total sample volume was greater than recommended, the lysate was divided into two AllPrep DNA Mini spin columns. From this point on, muscle and WAT samples were processed identically to the manufacturer’s instructions. RNA was stored at −80 °C until use.

Complementary DNA (cDNA) was synthesized from 0.5 µg of muscle tissue and myoblast RNA using QuantiTect Reverse Transcription Kit (Qiagen) according to the manufacturer’s instructions. In turn, SuperScript VILO cDNA Synthesis Kit (Thermofisher Scientific) was used to synthesize cDNA from 0.5 µg of WAT RNA, according to the manufacturer’s instructions.

Expression levels of genes associated with NAD^+^ biosynthesis, mitochondrial metabolism, mitochondrial OXPHOS, and additional potential NR target genes were determined from tissue samples with SYBRGreen-based quantitative reverse transcription PCR (qRT-PCR). Additionally, gene expression levels of myogenic marker *MYOG* and stem cell marker *PAX7* were measured from myoblasts in order to assess their differentiation potential. The PCR primers for these genes are provided in the Key resources table. qRT-PCR was performed using 1X SYBRGreen MasterMix (Thermofisher Scientific), 10 µM of primers and 5 ng of DNA template in 384 well plate format using CFX384 Touch™ Real-Time PCR Detection Systems (Bio-Rad Laboratories). Samples were run in duplicates with conditions otherwise identical to mtDNA quantification PCR. To normalize the expression data, three reference genes were used: actin, beta-2-microglobulin and tyrosine 3-monooxygenase/tryptophan 5-monooxygenase activation protein zeta. Data analysis was performed with the ΔΔCt method using the qBASE+ 3.2 software (Biogazelle).

### DNA methylation analysis

High molecular weight DNA was extracted from WAT and muscle using QiAmp DNA Mini kit (Qiagen) and bisulfite converted by EZ DNA Methylation kit (ZYMO Research) following the manufacturer’s protocol. DNA methylation was quantified using Infinium MethylationEPIC BeadChip arrays (Illumina) according to the manufacturer’s instructions. The data was preprocessed using R-package *minfi* (Aryee et al., 2014). Sample quality control was performed in R-package *MethylAid* with default settings (van Iterson et al., 2014). Furthermore, bad quality probes were removed with the following criteria: (i) detection p-value > 0.01, (ii) bead counts < 3, (iii) zero methylation probe intensity and (iv) ambiguously mapping probes identified by Zhou and colleagues (Zhou et al., 2017). The data were normalized using functional normalization (Fortin et al., 2014) followed by Beta Mixture Quantile normalization (Teschendorff et al., 2013). M values representing the methylation status of each probe were calculated by taking the log2 ratio of methylated probe intensity to unmethylated probe intensity. CpG sites that annotated to genes from which there was gene expression data available in either of the tissues, were selected for the analysis, resulting in a final dataset of 518 probes in WAT and 619 in muscle. Names of the genes are listed in the Key resources table.

Methylation levels in DNA repetitive elements (*e.g.* LINE-1) across the genome can be used as a surrogate measure for the global DNA methylation level of a sample. These elements are often hypermethylated, in contrast to CpGs located in other genomic areas (Rollins et al., 2006). The average LINE-1 methylation levels were computed using R-package *REMP* (Zheng et al., 2017) for each individual. Paired Wilcoxon signed-rank test was applied to compare the global methylation levels pre versus post NR across all twins from the BMI-discordant pairs (regardless of heavy/lean status), as well to compare global methylation levels as a result of NR supplementation, *i.e.* delta variables (change from the baseline to 5 months). The level of significance (two-tailed) was set at p<0.05.

Differential methylation analysis was performed for the selected CpGs in WAT (n=14 twin pairs / 28 individuals) and muscle (n=12 twin pairs / 25 individuals) using a linear model (package *limma* in R-Bioconductor) to identify those CpG sites that showed significantly different methylation levels (Benjamini-Hochberg FDR<0.05) pre versus post NR intervention across all twins from the BMI-discordant pairs (regardless of heavy/lean status). Regression analysis was also performed to identify CpGs that had different methylation levels as a result of NR intervention, *i.e.* delta variables (change from the baseline to 5 months). The regression models were adjusted for leaner/heavier status, sex, age, smoking status and twinship.

### Plasma metabolite isolation and non-targeted metabolite profiling

We performed, at the LC-MS metabolomics center (Biocenter Kuopio, University of Eastern Finland), non-targeted metabolite profiling on plasma metabolites. We analyzed samples using two different chromatographic-mass spectrometric techniques, which were reversed-phase (RP) and hydrophilic interaction (HILIC) chromatography techniques coupled online to high-resolution mass spectrometry, and further acquired data with positive and negative electrospray ionization (ESI) mode (Noerman et al., 2021; Pekkinen et al., 2013).

Briefly, plasma samples were thawed on ice and a 100 μl aliquot of plasma was dispensed into a 96-well Captiva ND filter plate 0.2 µm PP (Captiva ND, 0.2 µm PP, Agilent Technologies) containing 400 µl of ice-cold acetonitrile. Samples were mixed with a pipette to thoroughly precipitate plasma proteins. Samples were then centrifuged at 700 × g for 5 min at 4°C and the supernatants were collected to a 96-well storage plate and stored at −10°C.

A pooled sample from all biological samples per experiment was injected at the beginning of the sequence to equilibrate the analytical platform and then after every 12 samples throughout the analysis for quality control (QC). In addition, a solvent blank was prepared and injected at the beginning of the sequence. Samples were randomized before the analysis.

The analysis of amphiphilic metabolites was carried out using an ultra-high performance liquid chromatography (Vanquish Flex UHPLC system, Thermo Scientific, Bremen, Germany) coupled online to high-resolution mass spectrometry (HRMS, Q Exactive Classic, Thermo Scientific, Bremen, Germany). Samples were analyzed using the RP technique. The sample solution (2 µl) was injected onto an RP column (Zorbax Eclipse XDBC18, 2.1 × 100mm, 1.8 μm, Agilent Technologies, Palo Alto, CA, USA) that was kept at 40°C. The mobile phase, delivered at 400 μl/min, consisted of water (eluent A) and methanol (eluent B), both containing 0.1 % (v/v) of formic acid. The following gradient profile was used: 0–10 min: 2 to 100% B, 10–14.50 min: 100% B, 14.50–14.51 min: 100 to 2% B; 14.51–20 min: 2% B. The sample tray was at 10°C during these analyses. Mass spectrometry was equipped with heated electrospray ionization (ESI). The positive and negative ionization modes were used to acquire the data in centroid mode. At the beginning and end of the sample analysis, data-dependent product ion scans (MS2) were acquired for each mode. The detector was calibrated before the sample sequence and subsequently operated at high mass accuracy (<2 ppm). Continuous mass axis calibration was performed by monitoring reference ions m/z 214.08963 in the positive ionization mode.

For the analysis of hydrophilic compounds, an ultra-high performance liquid chromatography (1290 LC system, Agilent Technologies, Waldbronn, Karlsruhe, Germany) coupled online to high-resolution mass spectrometry (HRMS, 6540 UHD accurate-mass quadrupole-time-of-flight (qTOF) mass spectrometry, Agilent Technologies, Waldbronn, Karlsruhe, Germany) was used (Pekkinen et al., 2013). The sample solution (2 µl) was injected onto a column (HILIC, Acquity UPLC® BEH Amide 1.7 µm, 2.1×100 mm, Waters Corporation, Milford, MA, USA) that was kept at 45°C. Mobile phases, delivered at 600 μl/min, consisted of 50% (v/v)(eluent A) and 90% (v/v)(eluent B) acetonitrile, respectively, both containing 20 mM ammonium formate (pH 3). Following gradient profile was used: 0– 2.5 min: 100% B, 2.5–10 min: 100% B → 0% B, 10–10.1 min: 0% B → 100% B; 10.1–12.5 min: 100% B. The total runtime was 12.5 min. The sample tray was at 10°C during these analyses. Mass spectrometry was equipped with a heated ESI source, operated in both positive and negative ionization modes. For data acquisition, a 2 GHz extended dynamic range mode was used in both positive and negative ion modes from 50 to 1600 (m/z). Data was collected in the centroid mode. At the beginning and end of the sample analysis, data-dependent product ion scans (MS2) were acquired for each mode. The detector was calibrated before the sample sequence and subsequently operated at high accuracy (<2 ppm). Continuous mass axis calibration was performed by monitoring two reference ions from an infusion solution throughout the runs. The reference ions were *m/z* 121.050873 and *m/z* 922.009798 in the positive mode and *m/z* 112.985587 and *m/z* 966.000725 in the negative mode.

Further information about the instruments set-up and data acquisition parameters, in concert with molecular feature finding and peek picking can be obtained from the previous publications (Noerman et al., 2021; Pekkinen et al., 2013).

### Differential global metabolite profile analysis and metabolite identification

Differential metabolite analysis was performed on the global metabolite levels of the twins from BMI-discordant pairs (n=14 twin pairs/28 individuals) using a linear model (package *limma* in R-Bioconductor) to identify metabolite features that had significantly different levels (nominal p<0.05) pre vs post NR across all study subjects (regardless of heavy/lean status). Regression models were adjusted for leaner/heavier status, sex, age, smoking status and twinship (shared genetic effects). Regression analysis was also carried out to identify metabolite features showing different levels in heavier compared to leaner individuals as a result of NR intervention, *i.e.* delta metabolites (change from the baseline to 5 months). The regression models were adjusted for sex, age, smoking status and twinship. Significant metabolites were then further annotated, and enriched pathways were identified using MetaboAnalyst (metaboanalyst.ca) and Ingenuity Pathway Analysis tool (IPA, Qiagen).

The final data set contained 580 and 2398 molecular features from HILIC and RP analysis, respectively. Annotation of these significantly different metabolites found between the study groups was generated based on accurate mass and isotope information; *i.e.*, ratios, abundances, and spacing, as well as product ion spectra (MS2) against existing libraries, either in-house for level I or online spectral databases (*i.e.*, Human Metabolome Database, MzCloud, METLIN, ChemSpider) for level II-IV, according to the guidelines from the Metabolomics Standard Initiative (Sumner et al., 2007).

### Gut microbiota sequencing and composition analyses

Fecal samples were taken at home or at the hospital. For stool collection at home, study subjects were provided with the supplies and instructions for the collection. Study subjects were asked to keep the sample tube in a cold environment, and to bring the sample to the hospital. The stool was then immediately stored at −80 °C without any treatment. The interval between the passage of stool and storage at −80 °C was within 12 to 24 hours. The sample collection at the hospital was conducted in the same fashion. For the analyses of gut microbiota, the exclusion criteria were antibiotic course one month prior to the sample collection.

For the microbial community analysis, the total microbial DNA was extracted from ~80 mg of feces by first homogenizing the sample with a bead-beating method and then using Stool Extraction Kit v2 (Hain Lifescience GmbH, Nehren, Germany) and semi-automated GenoXtract (Hain Lifescience Gmbh). Thereafter, the rRNA gene was amplified using universal primers S-D-Bact-0341-b-S-17 and S-D-Bact-0785-a-A-21 targeting the V3-V4 regions of the SSU (small subunit) rRNA gene, details provided in the Key resource table. The PCR was performed using 1X SYBRGreen MasterMix (Thermofisher Scientific, Waltham, MA, USA), 0.5 µM of primers and 20 ng of DNA template. Thermal cycling for the first PCR was run using initial denaturation of 10 min at 95°C, 30 cycles of 30 s at 94°C and 60 s at 52°C, 60 s at 72°C and final extension for 5 min at 72°C. To add Ion Torrent PGM sequencing adapters and barcodes to the ends of the PCR product, one µl of the PCR product was used as a template in the second PCR, where 10 cycles were performed using linker and fusion primers (0.05 µM of M13_ S-D-Bact-0341-b-S-17, 0.5 µM of IonA_IonXpressBarcode_M13 and 0.5 µM P1_ S-D-Bact-0785-a-A-21), with conditions otherwise identical to the first amplification. Sequencing was performed using Ion Torrent PGM (Thermofisher Scientific). Prior to sequencing, the PCR products were purified with AMPure XP (Beckman Coulter), quantified with PicoGreen (Quant-iT PicoGreen dsDNA Assay Kit, Thermofisher Scientific) according to manufacturer’s instructions, and pooled in equimolar quantities for sequencing on Ion Torrent PGM using Thermofisher Scientific’s Hi-Q View OT2 Kit for emulsion PCR, Hi-Q View Sequencing Kit for the sequencing reaction, and Ion 318 Chip v2. The 16S rRNA gene sequences were quality-filtered and clustered to operational taxonomic units (OTUs) at 97% similarity using the CLC Microbial Genomics Package (Qiagen).

The rRNA gene sequences were classified using the SILVA SSU Ref database (99%, arb-silva.de/projects/ssu-ref-nr/). Statistical analyses for gut microbiota were performed with the CLC Microbial Genomics Package and SPSS Statistics (IBM). The alpha-diversity of the gut microbiota was quantified with the Shannon index and beta-diversity analysis was based on Bray-Curtis distance and PERMANOVA for significance testing. The taxonomic differences between pre versus post NR supplementation samples were analyzed with paired Wilcoxon signed-rank test in SPSS, followed by Benjamini-Hochberg FDR correction for multiple testing. The level of significance (two-tailed) was set at p<0.05 after the multiple testing correction.

Microbiota data were pre-processed and normalized using packages *phyloseq* and *edgeR* in R-Bioconductor. Differential analysis was performed for the gut microbiota levels of the twins (n=11 twin pairs/22 individuals) using a linear model (package *limma* in R-Bioconductor (Ritchie et al., 2015) to identify microbiota that had significantly different levels (nominal p<0.05) pre vs post NR across all study subjects (regardless of heavy/lean status). The regression models were adjusted for leaner/heavier status, sex, age, smoking status and twinship. Regression analysis was also carried out to identify differentially abundant microbiota showing different levels in heavier compared to leaner individuals as a result of NR intervention, *i.e.* delta microbiota (change from the baseline to 5 months). The regression models were adjusted for sex, age, smoking status and twinship.

*F. prausnitzii* qPCR analysis was performed using iQ SYBR Supermix and CFX96™ Real-time PCR Detection System (Bio-Rad Laboratories, Hercules, CA, USA). To amplify *F. prausnitzii,* 20 ng of template DNA and 10 pM of primers were used whereas to amplify the 16S rRNA gene, 10 ng of template DNA and 2.5 µM of primers were added per reaction. The PCR primers are provided in the Key resources table. The qPCR program for *F. prausnitzii* was as follows: pre-incubation at 95°C for 10 min, 40 cycles at 95°C for 30 sec, 60°C for 30 sec and 72°C for 15 sec. The qPCR program for the 16S rRNA genomic region was pre-incubation at 95°C for 3 min, 40 cycles at 95°C for 30 sec, 53°C for 30 sec and 72°C for 30 sec. The results of *F. prausnitzii* were normalized to the results of 16S rRNA, and the fold change between the pre and post NR samples was calculated with the ΔΔCt method. Due to a great interindividual variation, the effect of NR on *F. prausnitzii* amount was analyzed with Cohen’s d as follows:

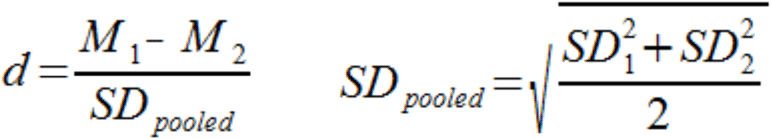

where *M_1_* and *M_2_* are the means for the 1st and 2nd samples, and *SD_pooled_* is the pooled standard deviation for the samples.

### Association analyses

Associations between CpG methylation (outcome variable) and gene expression (explanatory variables) were identified in study participants with the respective data available. First, delta gene expression (the change from baseline to 5 months) was calculated which was then compared to corresponding delta DNA methylation (the change from baseline to 5 months) of the given gene. Linear modelling was used to test for the associations, adjusting for age, sex, smoking status and twinship. Benjamini-Hochberg FDR correction was performed separately for each CpG-variable comparison with a significance threshold of <0.05.

Furthermore, the associations between NAD/global metabolites (outcome variable) and microbiota (explanatory variables) were examined. First, regression analysis was performed to identify changes, between 0 and 5 months, in NAD metabolites in relation to changes in the microbiota. Next, we identified, between 0 and 5 months, changes in NAD metabolites in relation to the baseline microbiota. Finally, a similar analysis was performed again, to identify changes, between 0 and 5 months, in global metabolites as well as clinical parameters, in relation to changes in the microbiota. Since our focus was on the metabolites that had already been determined to be differentially abundant in pre vs post NR samples, only their relationships with microbiota were examined. For relationships, *i.e.* associations, that were significant (FDR<0.05), pathway analysis was performed using the IPA tool. All of the regression models mentioned above were adjusted for leaner/heavier status, sex, age, antibiotics usage, smoking status and twinship.

## Quantification and statistical analysis

Clinical variables, blood NAD metabolites, mitochondria and gene expression data are expressed as mean ± standard deviation (SD) for variables with normal distribution, and as median (interquartile range) for variables with non-normal distribution. The normality of the distribution was determined using the Shapiro-Wilk test. Variables with a p-value of less than 0.05 were considered non-normally distributed. Outlier removal for gene expression and mtDNA amount data was performed in GraphPad Prism by using outlier analysis ROUT (Q=1). Differences between the cotwins from the BMI-discordant and BMI-concordant pairs at baseline, pre versus post treatment across all study participants and delta variables between the cotwins from the BMI-discordant pairs were assessed using the paired Wilcoxon signed-rank test using R software and the level of significance (two-tailed) was set at p<0.05. In the figure and table legends, we state the specific statistical tests used as well as the number of included twin pairs/individuals, and the cutoffs for statistical significance.

## Additional resources

Clinicaltrials.gov identifier NCT03951285.

