## Supplemental figures and tables 1-4 and 10-11 for "Nicotinamide riboside improves muscle mitochondrial biogenesis, satellite cell differentiation and gut microbiota composition in a twin study"

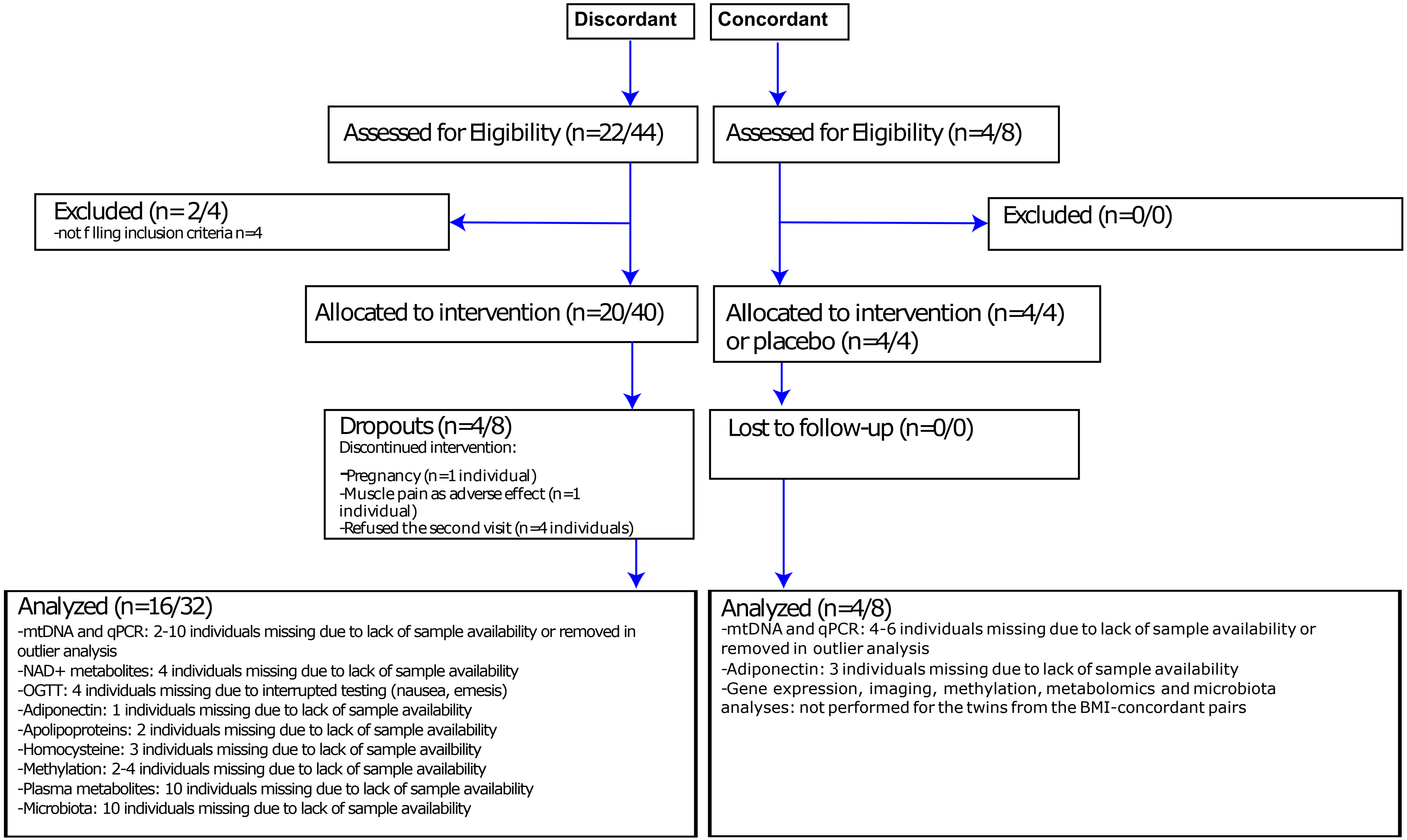


**Fig. S1. Flow chart illustrating the procedures for selection of study participants and data analyses, Related to Figure 1A. Presented as (n=number of twin pairs/number of individuals).**

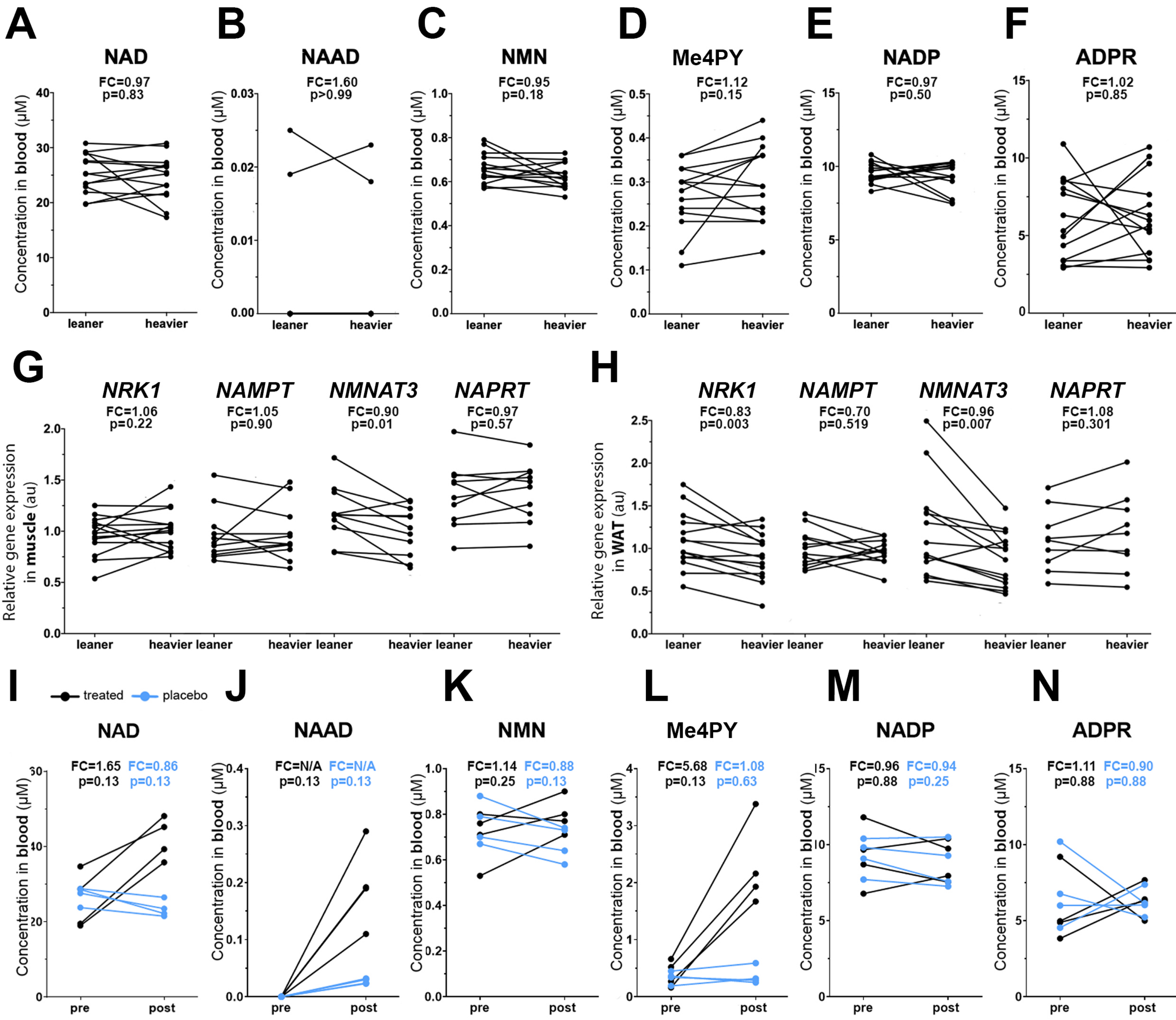


**Fig. S2. Baseline NAD+ metabolism in the twins from the BMI-discordant pairs and the effect of placebo and NR treatments on NAD metabolome in the twins from the BMI-concordant pairs. Related to Figure 1. Number of analyzed study participants is presented as n=number of full twin pairs/number of individuals.**

(A-F) Concentration of whole blood NAD metabolites at baseline in the twins from the BMI-discordant pairs (n=14 pairs/28 individuals).

(G) Expression of genes associated with NAD+ biosynthesis in muscle at baseline in the twins from the BMI-discordant pairs (n=11-15 pairs/22-30 individuals).

(H) Expression of genes associated with NAD+ biosynthesis in WAT at baseline in the twins from the BMI-discordant pairs (n=9-14 pairs/18-28 individuals).

(I-N) Concentration of whole blood NAD metabolites in placebo and NR treated BMI-concordant twin pairs (n=4 pairs/8 individuals) .One cotwin of the twin pair was treated with NR (in black) and the other cotwin with placebo (in blue).

Pre and post values of an individual study subject are connected with a line. Fold change (FC) indicates either the mean of the heavier cotwin values divided with the leaner cotwin values (baseline graphs) or post NR values divided by pre values (pre versus post graphs). FC could not be calculated for NAAD as its concentration at baseline was zero. Arbitrary unit (au) indicates relative gene expression normalized to expression of housekeeping genes. Paired Wilcoxon signed-rank test was used as the statistical test. NAAD, nicotinic acid adenine dinucleotide; NMN,nicotinamide mononucleotide; Me4PY, N-methyl-4-pyridone-5-carboxamide; NADP, nicotinamide adenine dinucleotide phosphate; ADPR, ADP ribose.

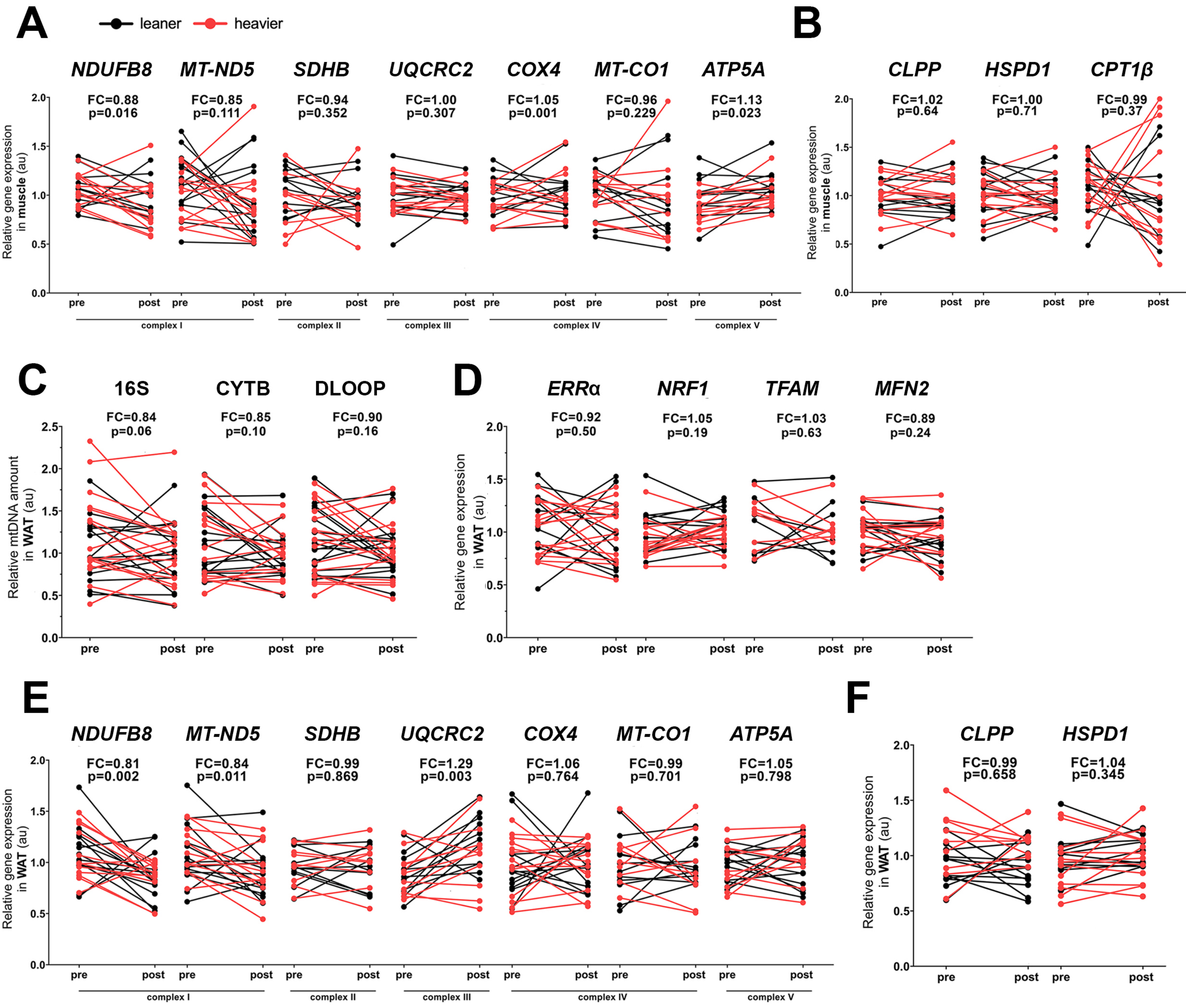


**Fig. S3. Expression of mitochondrial metabolism related genes in muscle and WAT and WAT mitochondrial DNA amount before and after NR treatment in the twins from the BMI-discordant pairs. Related to Figure 2. Number of analyzed study participants is presented as n=number of full twin pairs/number of individuals.**

(A) Gene expression of OXPHOS complex (I-V) subunits in muscle pre versus post NR (n= 10-13 pairs/21-26 individuals).

(B) Expression of mitochondrial quality control and fatty acid oxidation genes in muscle pre versus post NR (n= 11-13 pairs/23-26 individuals).

(C) Mitochondrial DNA amount in WAT pre versus post NR (n=14-15 pairs/28-31 individuals).

(D) Gene expression levels of the regulators of mitochondrial biogenesis in WAT pre versus post NR (n=10-13 pairs/21-27 individuals)

(E) Gene expression levels of OXPHOS complex (I-V) subunits in WAT pre vs post NR (n=9-13 pairs/19-26 individuals).

(F) Expression levels of mitochondrial quality control genes in WAT pre vs post NR (n= 11 pairs/22-23 individuals).

Pre and post values of an individual study subject are connected with a line and leaner twins are in black, heavier in red.

Fold change (FC) indicates either mean of post NR values divided by pre values. Paired Wilcoxon signed-rank test was used as the statistical test. Arbitrary unit (au) indicates relative gene expression normalized to expression of housekeeping genes or mitochondrial amount expressed relative to nuclear genome amount. mtDNA, mitochondrial amount.

**Table S1. Baseline characteristics of the twins from the BMI-discordant and -concordant pairs.**

| Variable (unit) | Discordant All (*n*=32 individuals) | Leaner (*n*=16 individuals) | Heavier (*n*=16 individuals) | P Value | Concordant All (*n*=8 individuals) | Concordant Placebo (*n*=4 individuals) | Concordant Treated (*n*=4 individuals) | P Value |
|---|---|---|---|---|---|---|---|---|
| Age (years) | 39.7 (33.1 - 41.1) | 39.7 (33.1 - 41.1) | 39.7 (33.1 - 41.1) | - | 40.5 (37.8 - 41.3) | 40.5 (37.8 - 41.3) | 40.5 (37.8 - 41.3) | - |
| Sex (female, %) | 44 | 44 | 44 | - | 50 | 50 | 50 | - |
| Height (cm) | 173.4 ± 10.9 | 173.5 ± 11.5 | 173.3 ± 10.7 | 0.477 | 167.4 ± 10.1 | 167.4 ± 11.1 | 167.4 ± 10.8 | 0.875 |
| BMI ($kg/m^2$) | 30.1 ± 5.7 | 27.4 ± 4.2 | 32.8 ± 5.8 | <0.001 | 31.7 ± 6.4 | 31.5 ± 6.8 | 32.0 ± 6.9 | 0.269 |
| Weight (kg) | 90.7 ± 19.8 | 82.7 ± 17.2 | 98.6 ± 19.5 | <0.001 | 88.1 ± 13.5 | 87.2 ± 12.2 | 89.1 ± 16.6 | 0.625 |
| Waist-hip ratio | 0.96 ± 0.09 | 0.9 ± 0.1 | 1.0 ± 0.1 | <0.001 | 0.94 ± 0.04 | 0.9 ± 0.0 | 1.0 ± 0.0 | 0.250 |
| Body fat (%) | 37.1 ± 8.2 | 34.0 ± 7.8 | 40.2 ± 7.6 | <0.001 | 33.4 ± 12.8 | 34.1 ± 13.8 | 33.1 ± 14.8 | 1.000 |
| Visceral adipose tissue ($cm^3$) | 2155 ± 878 | 1754 ± 786 | 2556 ± 799 | 0.001 | 2117 ± 693 | 2017 ± 497 | 2217 ± 921 | 0.875 |
| Subcutaneous adipose tissue ($cm^3$) | 6461 ± 3001 | 5325 ± 2442 | 7597 ± 3155 | <0.001 | 7146 ± 4461 | 7297 ± 4799 | 6996 ± 4831 | 0.250 |
| Adipocyte cell number (per billion) | 301 (194 - 505) | 288 (173 - 383) | 357 (203 - 581) | 0.010 | 191 (173 - 267) | 233 (197 - 308) | 168 (147 - 204) | 0.125 |
| Adipocyte cell diameter (µM) | 93 ± 11 | 88 ± 10 | 97 ± 8.9 | 0.002 | 93 ± 12 | 89 ± 11 | 96 ± 12 | 0.250 |
| Adipocyte cell volume (pL) | 598 ± 201 | 508 ± 213 | 689 ±145 | 0.007 | 559 ± 186 | 500 ± 154 | 617 ± 219 | 0.250 |
| Adipocyte cell weight (ng) | 583 (429 - 641) | 435 (306 - 572) | 627 (583 - 740) | 0.007 | 452 (365 - 674) | 407 (364 - 500) | 584 (427 - 722) | 0.250 |
| PPARγ, adipose tissue (au) | 0.95 ± 0.30 | 1.00 ± 0.34 | 0.91 ± 0.25 | 0.569 | ND | ND | ND | ND |
| Liver fat (%) | 1.7 (0.8 - 4.6) | 1.2 (0.5 - 2.0) | 2.3 (1.4 - 9.2) | 0.010 | 3.4 (1.2 - 8.7) | 5.2 (0.88 - 9.9) | 3.4 (2.0 - 5.4) | 0.625 |
| Lean tissue mass (kg) | 54.0 ± 10.2 | 52.0 ± 9.6 | 56.1 ± 10.5 | <0.001 | 57.5 ± 4.9 | 56.2 ± 5.5 | 58.8 ± 4.9 | 0.250 |
| Total bone mass (kg) | 2.9 ± 0.6 | 2.8 ± 0.6 | 2.9 ± 0.6 | <0.001 | 3.2 ± 0.5 | 3.2 ± 0.6 | 3.3 ± 0.5 | 0.500 |
| Basal metabolic rate (kcal/day) | 1874 ± 382 | 1780.8 ± 370.4 | 1967.7 ± 380.9 | <0.001 | 1797.9 ± 262.1 | 1782.3 ± 265.8 | 1813.5 ± 298.3 | 0.625 |
| Caloric intake (kcal/day) | 2297 ± 441 | 2244.3 ± 434.4 | 2349.9 ± 455.7 | 0.463 | 2003.1 ± 655.0 | 1872.0 ± 419.2 | 2134.2 ± 882.8 | 1.000 |

| | | | | | | | | |
|---|---|---|---|---|---|---|---|---|
| Protein intake (g) | 96.2 (81.6 - 115) | 95.6 (84.5 - 115.3) | 96.3 (79.2 - 116.4) | 0.900 | 81.8 (65.3 - 103.5) | 81.5 (6.2 - 100.2) | 84.0 (65.7 - 110.1) | 0.625 |
| Fat intake (g) | 103.6 ± 28.5 | 97.7 ± 26.0 | 109.6 ± 30.4 | 0.562 | 78.4 ± 29.7 | 69.8 ± 17.5 | 87.1 ± 39.5 | 0.875 |
| Carbohydrate intake (g) | 211.5 ± 51.1 | 216.0 ± 50.0 | 207.0 ± 53.4 | 0.744 | 212.4 ± 68.2 | 234.9 ± 55.1 | 221.3 ± 93.2 | 0.875 |
| Niacin equivalent (mg/day) | 44.5 (36.1 - 49.9) | 45.4 (36.7 - 49.3) | 43.9 (33.5 - 52.9) | 0.706 | 33.0 (31.0 - 40.7) | 32.4 (30.6 - 36.5) | 35.8 (31.9 - 41.6) | 0.875 |
| Physical activity (Total Baecke) | 7.8 ± 1.5 | 8.0 ± 1.7 | 7.7 ± 1.4 | 0.802 | 8.4 ± 1.4 | 8.1 ± 0.7 | 8.7 ± 1.9 | 0.750 |
| Alcohol consumption (doses/week) | 1.0 (0.0 - 2.0) | 0.9 (0 - 3.8) | 5.0 (1.8 - 9.3) | 0.009 | 1.0 (0.0 - 2.9) | 1.0 (0.5 - 1.4) | 2.0 (0.0 - 4.6) | 0.371 |
| Current smoker (number) | 7 | 4 | 3 | - | 3 | 2 | 1 | - |
| HbA1c (mmol/mol) | 34 (32 - 36) | 34 (32 - 35) | 35 (34 - 36) | 0.037 | 36 (33 - 37) | 35 (33 – 36) | 37 (35 – 38) | 1.000 |
| Fasting glucose (mmol/l) | 5.6 (5.4 - 5.8) | 5.6 (5.3 - 5.7) | 5.7 (5.4 - 5.8) | 0.157 | 5.8 (5.1 - 5.8) | 5.4 (5.0 - 5.8) | 5.8 (5.6 - 5.8) | 0.375 |
| Fasting insulin (mlU/l) | 6.0 (4.5 - 9.8) | 4.7 (3.8 - 6.7) | 7.9 (5.3 - 12.5) | 0.018 | 8.0 (3.9 - 12.9) | 7.7 (3.9 - 11.5) | 9.7 (4.1 - 14.9) | 0.375 |
| Fasting C-peptide (nmol/l) | 0.51 (0.39 - 0.65) | 0.47 (0.39 - 0.60) | 0.56 (0.43 - 0.83) | 0.018 | 0.69 (0.44 - 1.1) | 0.69 (0.41 - 0.98) | 0.80 (0.44 - 1.2) | 0.125 |
| HOMA index | 1.5 (1.0 - 2.1) | 1.1 (0.94 - 1.6) | 2.0 (1.3 - 3.2) | 0.022 | 1.9 (0.96 - 3.4) | 1.8 (0.96 - 2.7) | 2.5 (1.0 - 3.8) | 0.250 |
| Matsuda index | 5.7 ± 2.5 | 6.8 ± 2.4 | 4.7 ± 2.1 | <0.001 | 8.2 ± 6.2 | 8.8 ± 6.9 | 7.5 ± 6.5 | 0.125 |
| Glucose AUC during OGTT | 15.7 ± 2.5 | 15.2 ± 2.5 | 16.2 ± 2.4 | 0.042 | 15.8 ± 3.6 | 14.8 ± 4.1 | 16.8 ± 3.3 | 0.125 |
| Insulin AUC during OGTT | 90 (72 - 135) | 85.3 (67.7 - 116.9) | 96.6 (78.4 - 175.2) | 0.006 | 95.5 (30.9 - 188.0) | 90.0 (30.3 - 161.3) | 115.2 (39.2 - 188.0) | 0.625 |
| Adiponectin (µg/ml) | 3.2 (2.1 - 4.3) | 3.2 (2.7 - 3.8) | 3.1 (1.6 - 5.2) | 0.454 | 3.4 (2.2 - 8.5) | 3.4 (2.8 - 6.8) | 5.2 (3.6 - 6.9) | 0.500 |
| Cholesterol (total, mmol/l) | 4.7 ± 0.8 | 4.7 ± 0.7 | 4.7 ± 0.9 | 0.660 | 4.8 ± 0.6 | 4.7 ± 0.5 | 4.8 ± 0.8 | 0.875 |
| HDL (mmol/l) | 1.3 (1.2 - 1.6) | 1.4 (1.2 - 1.5) | 1.3 (1.1 - 1.7) | 0.224 | 1.3 (1.2 - 1.5) | 1.3 (1.2 - 1.4) | 1.4 (1.2 - 1.6) | 0.625 |
| LDL (mmol/l) | 3.0 ± 0.77 | 3.0 ± 0.71 | 3.1 ± 0.83 | 0.196 | 3.2 ± 0.41 | 3.1 ± 0.41 | 3.3 ± 0.45 | 0.586 |
| Triglycerides (mmol/l) | 0.95 (0.71 - 1.2) | 0.81 (0.61 - 1.0) | 1.0 (0.73 - 1.3) | 0.118 | 1.0 (0.70 - 1.1) | 1.0 (0.88 - 1.0) | 0.92 (0.70 - 1.2) | 0.625 |
| FFA (µmol/l) | 444 (376 – 551) | 415 (362 - 500) | 510 (428 - 700) | 0.542 | 398 (332 - 527) | 427.7 (325.4 - 531.4) | 394.1 (332.1 - 481.9) | 0.625 |

| | | | | | | | | |
|---|---|---|---|---|---|---|---|---|
| Systolic BP (mmHg) | 133 ± 22 | 131 ± 24 | 135 ± 22 | 0.141 | 120 ± 12 | 114 ± 12 | 126 ± 7.8 | 0.098 |
| Diastolic BP (mmHg) | 83 ± 12 | 83 ± 12 | 84 ± 11 | 0.706 | 77 ± 8 | 77 ± 7.7 | 80 ± 5.9 | 0.197 |
| Pulse (per min) | 66.9 ± 10.7 | 64.5 ± 9.2 | 69.3 ± 11.8 | 0.084 | 61.9 ± 6.4 | 63.0 ± 8.0 | 60.8 ± 5.1 | 0.461 |
| Hs-CRP (mg/l) | 1.9 (0.67 - 3.9) | 1.7 (0.61 - 3.0) | 2.3 (0.73 - 5.8)) | 0.117 | 1.3 (0.83 - 5.4) | 1.6 (1.3 - 10.7) | 0.99 (0.55 - 3.3) | 0.250 |
| Total homocysteine (μmol/l) | 11.0 (9.30 -12.0) | 10.0 (9.55 -12.0) | 11.0 (9.28 -12.8) | 0.500 | 13.0 (11.5 - 13.8) | 13.0 (12.2 - 13.8) | 12.5 (11.5 - 15.0) | 1.000 |
| ALT (U/l) | 22.0 (17.8 - 42.8) | 20.0 (16.5 - 33.5) | 33.0 (19.5 - 49.0) | 0.012 | 22.5 (19.8 - 32.0) | 23.5 (18.3 - 38.8) | 22.5 (21.5 - 28.3) | 0.581 |
| AST (U/l) | 24.0 (21.8 - 31.0) | 24.0 (19.8 - 29.2) | 24.0 (22.8 - 31.0) | 0.139 | 26.5 (24.0 - 29.5) | 26.5 (24.8 - 30.0) | 26.5 (24.0 - 29.5) | 0.586 |
| Creatinine (μmol/l) | 73.7 ± 12.9 | 73.2 ± 13.6 | 74.3 ± 12.7 | 0.569 | 76.6 ± 12.0 | 74.0 ± 13.2 | 79.3 ± 12.1 | 0.125 |
| Haemoglobin (g/l) | 142 ± 9 | 142 ± 8 | 143 ± 9 | 0.659 | 136 ± 11 | 137 ± 10 | 136 ± 13 | 0.625 |
| Haematocrit (%) | 42.0 ± 2.2 | 41.9 ± 2.1 | 42.1 ± 2.4 | 0.680 | 40.6 ± 3.2 | 40.8 ± 2.9 | 40.5 ± 3.9 | 0.850 |
| Erythrocytes ($x10^{12}$/l) | 4.8 ± 0.3 | 4.8 ± 0.37 | 4.8 ± 0.31 | 0.103 | 4.5 ± 0.4 | 4.5 ± 0.4 | 4.6 ± 0.5 | 0.875 |
| MCV (fl) | 87.9 ± 3.5 | 88.4 ± 4.2 | 87.4 ± 2.6 | 0.162 | 89.0 ± 3.3 | 90.3 ± 2.1 | 87.8 ± 4.2 | 0.098 |
| MCH (pg) | 29.8 ± 1.3 | 29.9 ± 1.5 | 29.6 ± 1.2 | 0.182 | 30.1 ± 1.4 | 30.5 ± 1.0 | 29.8 ± 1.7 | 0.371 |
| MCHC (g/l) | 338 (334 - 342) | 338 (334 - 342) | 337 (335 - 342) | 0.609 | 338 (334.5 - 335.3) | 338.0 (335.5 - 339.8) | 338.5 (334.5 - 342.5) | 0.345 |
| Leukocytes ($x10^9$/l) | 6.4 ± 1.5 | 6.5 ± 1.8 | 6.3 ± 1.2 | 0.816 | 6.7 ± 2.3 | 7.3 ± 3.0 | 6.2 ± 1.7 | 0.423 |
| Thrombocytes ($x10^9$/l) | 243 ± 55 | 244 ± 60 | 241 ± 52 | 0.860 | 247 ± 42 | 234 ± 29 | 260 ± 53 | 0.125 |
| fP-Fe (μmol/l) | 19.0 ± 6.9 | 19.8 ± 7.6 | 18.1 ± 6.3 | 0.469 | 20.0 ± 7.3 | 20.0 ± 6.4 | 20.0 ± 9.1 | 0.875 |

Data are shown as mean ± standard deviation or median (interquartile range). P values were obtained using paired Wilcoxon rank-sum test to examine the differences on anthropometric and clinical parameters between the cotwins at baseline in the twins from the BMI-discordant (n=16 twin pairs/32 individuals) and concordant (n=4 twin pairs/8 individuals) pairs. *PPAR*γ, peroxisome proliferator-activated receptor γ; Au, arbitrary unit; ND, not determined; HbA1c, hemoglobin A1c; HOMA, homeostasis model assessment; AUC, area under curve; OGTT, oral glucose tolerance test; HDL, high-density lipoprotein; LDL, low-density lipoprotein; FFA, free fatty acids; BP, blood pressure; Hs-CRP, high sensitive C-reactive protein; ALT, alanine aminotransferase; AST, aspartate transaminase; MCV, mean corpuscular volume; MCH, mean corpuscular hemoglobin; MCHC, mean corpuscular hemoglobin concentration; fP, fasting plasma and Fe, iron.

**Table S2. The delta response (the change from baseline to 5 months) to NR in the twins from the BMI-discordant pairs.**

| Variable (unit) | Leaner Δ (*n*=16 individuals) | Heavier Δ (*n*=16 individuals) | P Value |
|---|---|---|---|
| *NAD metabolites* | | | |
| $NAD^+$, whole blood (μM) | 30.1 ± 20.0 | 33.4 ± 12.1 | 0.519 |
| NAAD, whole blood (μM) | 0.15 (0.08 - 0.34) | 0.26 (0.11 - 0.31) | 0.533 |
| NMN, whole blood (μM) | 0.12 ± 0.15 | 0.23 ± 0.08 | 0.139 |
| NAR, whole blood (μM) | 0.0 (0.0 - 0.0) | 0.0 (0.0 - 0.0) | 1.000 |
| Me4py, whole blood (μM) | 1.6 ± 0.59 | 1.9 ± 0.46 | 0.014 |
| NADP, whole blood (μM) | -0.82 (-1.7 - -0.69) | -0.15 (-0.88 - 0.60) | 0.175 |
| ADPR, whole blood (μM) | -0.85 ± 3.1 | 0.42 ± 2.8 | 0.557 |
| *Expression of $NAD^+$ biosynthesis genes* | | | |
| *NRK1*, muscle (au) | 0.08 ± 0.14 | 0.04 ± 0.13 | 0.424 |
| *NAMPT*, muscle (au) | 0.13 (-0.04 - 0.29) | 0.19 (0.09 - 0.18) | 0.742 |
| *NMNAT3*, muscle (au) | -0.24 ± 0.27 | -0.06 ± 0.26 | 0.129 |
| *NAPRT*, muscle (au) | -0.10 ± 0.20 | -0.10 ± 0.21 | 1.000 |
| *NRK1*, adipose tissue (au) | 0.01 ± 0.28 | 0.19 ± 0.23 | 0.068 |
| *NAMPT*, adipose tissue (au) | 0.06 (-0.08 - 0.23) | -0.04 (-0.13 - 0.20) | 0.922 |
| *NMNAT3*, adipose tissue (au) | 0.01 (-0.25 - 0.24) | 0.09 (0.04 - 0.23) | 0.110 |
| *NAPRT*, adipose tissue (au) | -0.05 (-0.24 - 0.12) | -0.15 (-0.50 - -0.05) | 0.938 |
| *Anthropometric and body composition* | | | |
| BMI ($kg/m^2$) | 0.89 ± 1.5 | 1.0 ± 1.2 | 0.224 |
| Weight (kg) | 2.7 ± 4.3 | 2.6 ± 3.4 | 0.339 |
| Body fat (%) | 1.5 ± 2.6 | 0.62 ± 2.2 | 0.404 |
| Visceral adipose tissue ($cm^3$) | 250 ± 307 | 101 ± 560 | 0.610 |
| Subcutaneous adipose tissue ($cm^3$) | 683 ± 1240 | -46 ± 1295 | 1.000 |
| Adipose tissue ($cm^3$) | 933 ± 1476 | 55 ± 1760 | 1.000 |
| Adipocyte cell number (per billion) | -123 (-150 - -4.0) | -6.2 (-337 - 95) | 0.599 |
| Adipocyte cell diameter (μM) | 4.5 ± 16 | 1.0 ± 17 | 0.454 |
| Adipocyte cell volume (pL) | 111 ± 356 | 110 ± 17 | 0.639 |
| Adipocyte cell weight (ng) | -6.3 (-113 - 186) | 38 (-159 - 365) | 0.639 |
| PPARγ, adipose tissue (au) | 0.13 ± 0.26 | 0.15 ± 0.22 | 0.765 |
| Waist-hip ratio | 0.02 ± 0.05 | 0.00 ± 0.03 | 0.211 |
| Liver fat (%) | 0.05 (-0.25 - 0.55) | -0.10 (-0.70 - 1.10) | 0.635 |
| Lean tissue mass (g) | 158 ± 1234 | 747 ± 1322 | 0.193 |
| Total bone mass (g) | 10.5 ± 32.2 | 6.6 ± 17.4 | 0.940 |
| Basal metabolic rate (kcal/day) | 30.8 ± 54.0 | 47.0 ± 44.4 | 0.313 |
| Caloric intake (kcal/day) | 20 ± 627 | -135 ± 687 | 0.562 |
| Niacin equivalent (mg/day) | -4.0 (-12.5 - 2.1) | 0.47 (-14.7 - 11.2) | 0.528 |
| Physical activity (Total Baecke) | 0.0 ± 1.5 | -0.3 ± 0.6 | 0.401 |
| Alcohol intake (doses/week) | 0.49 (0.0 - 0.94) | -0.19 (-0.50 - 0.13) | 0.074 |
| *Glucose homeostasis* | | | |
| HbA1c (mmol/mol) | 1.0 (0.0 - 1.0) | 0.0 (-2.0 - 1.5) | 0.037 |
| Fasting glucose (mmol/l) | 0.10 (-0.10 - 0.20) | 0.20 (-0.10 - 0.33) | 0.089 |

| | | | |
|---|---|---|---|
| Fasting insulin (mlU/l) | 1.2 (-0.1 - 4.1) | 1.5 (-1.2 - 3.6) | 0.670 |
| Fasting C-peptide (nmol/l) | 0.05 (0.01 - 0.15) | 0.11 (-0.0 - 0.18) | 0.599 |
| HOMA index | 0.35 (-0.11 - 1.0) | 0.37 (-0.22 - 0.98) | 0.804 |
| Matsuda index | -1.4 ± 2.1 | -1.0 ± 1.8 | 0.305 |
| Glucose AUC during OGTT | 0.3 ± 2.2 | 0.6 ± 1.4 | 0.635 |
| Insulin AUC during OGTT | 14.8 (-8.8 - 30.7) | 11.0 (-1.6 - 64.8) | 0.635 |
| Adiponectin (ng/ml) | 17 (-111 - 516) | -105 (-796 - 364) | 0.151 |
| *Cardiovascular health* | | | |
| Cholesterol (total, mmol/l) | 0.03 ± 0.33 | 0.07 ± 0.32 | 0.889 |
| HDL (mmol/l) | -0.08 (-0.17 - 0.03) | -0.05 (-0.18 - 0.08) | 0.231 |
| LDL (mmol/l) | 0.03 ± 0.35 | -0.08 ± 0.59 | 0.551 |
| Triglycerides (mmol/l) | 0.13 (-0.11 - 0.26) | 0.15 (-0.08 - 0.27) | 0.231 |
| FFA (μmol/l) | -9.48 (-89.3 - 47.7) | 36.5 (-154 - 66.5) | 0.542 |
| Systolic BP (mmHg) | -4.0 ± 17.5 | -1.7 ± 17.5 | 0.615 |
| Diastolic BP (mmHg) | -0.5 ± 9.5 | 1.1 ± 13.9 | 0.624 |
| Pulse (per min) | 2.1 ± 9.1 | 0.14 ± 12.9 | 0.575 |
| Hs-CRP (mg/l) | 0.13 (-0.39 - 0.65) | -0.07 (-0.43 - 0.71) | 0.391 |
| Total homocysteine (μmol/l) | 2.0 (0.15 - 2.5) | 0.90 (-0.93 - 1.2) | 0.024 |
| *Mitochondrial parameters* | | | |
| Mitochondrial number, muscle (number/muscle fiber area/10 $\mu m^2$) | 12.6 ± 23.0 | 12.0 ± 29.9 | 1.000 |
| Mitochondrial area, muscle (relative area %)) | 0.67 (0.01 - 1.12) | 0.91 (-0.33 - 2.2) | 1.000 |
| Mitochondrial perimeter (μm) | -0.02 ± 0.28 | 0.04 ± 0.35 | 0.966 |
| Mitochondrial diameter (μm) | 0.01 (-0.02 - 0.02) | -0.02 (-0.02 - 0.02) | 0.413 |
| Mitochondrial form factor | -0.09 (-0.17 - 0.25) | 0.07 (-0.06 - 0.27) | 0.365 |
| Mitochondrial aspect ratio | -0.05 (-0.25 - 0.26) | 0.09 (-0.08 - 0.24) | 0.240 |
| 16S, muscle (au) | 0.27 (0.25 - 0.53) | 0.30 (0.01 - 0.68) | 0.846 |
| CYTB, muscle (au) | 0.31 (0.16 - 0.37) | 0.18 (0.11 - 0.47) | 0.622 |
| DLOOP, muscle (au) | 0.29 ± 0.16 | 0.31 ± 0.24 | 0.922 |
| 16S, adipose tissue (au) | -0.09 ± 0.42 | -0.19 ± 0.40 | 0.542 |
| CYTB, adipose tissue (au) | -0.06 (-0.52 - 0.06) | -0.04 (-0.34 - 0.08) | 0.635 |
| DLOOP, adipose tissue (au) | -0.11 ± 0.41 | -0.11 ± 0.34 | 0.934 |
| *ERRα*, muscle (au) | 0.15 ± 0.23 | 0.15 ± 0.19 | 0.413 |
| *NRF1*, muscle (au) | -0.08 ± 0.25 | -0.16 ± 0.32 | 0.547 |
| *PGC1α*, muscle (au) | -0.07 ± 0.35 | -0.04 ± 0.38 | 0.791 |
| *TFAM*, muscle (au) | 0.16 ± 0.34 | 0.28 ± 0.36 | 0.244 |
| *MFN2*, muscle (au) | 0.14 ± 0.15 | 0.06 ± 0.21 | 0.520 |
| *NDUFB8*, muscle (au) | -0.09 ± 0.25 | -0.19 ± 0.27 | 0.770 |
| *MT-ND5*, muscle (au) | -0.10 (-0.59 - 0.20) | -0.17 (-0.44 - 0.01) | 0.893 |
| *SDHB*, muscle (au) | -0.04 ± 0.24 | 0.12 ± 0.55 | 0.734 |
| *UQCRC2*, muscle (au) | 0.01 ± 0.19 | 0.02 ± 0.23 | 0.685 |
| *COX4,* muscle (au) | -0.07 (-0.18 - 0.16) | 0.05 (-0.14 - 0.26) | 0.938 |
| *MT-CO1*, muscle (au) | -0.06 ± 0.31 | -0.02 ± 0.42 | 0.820 |
| *ATP5A*, muscle (au) | 0.10 ± 0.22 | 0.14 ± 0.16 | 0.910 |
| *CLPP*, muscle (au) | -0.01 ± 0.17 | 0.04 ± 0.19 | 0.557 |

| | | | |
|---|---|---|---|
| *HSPD1*, muscle (au) | -0.04 ± 0.26 | 0.00 ± 0.25 | 0.733 |
| *CPT1β,* muscle (au) | -0.15 ± 0.63 | -0.03 ± 0.73 | 0.765 |
| *ERRα*, adipose tissue (au) | -0.07 ± 0.46 | -0.02 ± 0.27 | 0.339 |
| *NRF1*, adipose tissue (au) | 0.08 (-0.03 - 0.16) | 0.11 (-0.09 - 0.21) | 0.787 |
| *TFAM*, adipose tissue (au) | -0.02 ± 0.39 | -0.05 ± 0.26 | 1.000 |
| *MFN2*, adipose tissue (au) | -0.07 ± 0.21 | -0.05 ± 0.25 | 0.898 |
| *NDUFB8*, adipose tissue (au) | -0.30 (-0.52 - -0.07) | -0.36 (-0.42 - -0.11) | 0.588 |
| *MT-ND5*, adipose tissue (au) | -0.19 (-0.38 - 0.02) | -0.17 (-0.43 - 0.01) | 0.622 |
| *SDHB*, adipose tissue (au) | -0.05 (-0.25 - 0.09) | -0.07 (-0.13 - 0.11) | 0.742 |
| *UQCRC2*, adipose tissue (au) | 0.30 ± 0.38 | 0.16 ± 0.26 | 0.426 |
| *COX4*, adipose tissue (au) | 0.04 (-0.39 - 0.30) | -0.01 (-0.15 - 0.11) | 1.000 |
| *MT-CO1*, adipose tissue (au) | 0.02 (-0.14 - 0.11) | -0.13 (-0.30 - -0.17) | 0.734 |
| *ATP5A*, adipose tissue (au) | -0.04 ± 0.32 | 0.06 ± 0.23 | 0.365 |
| *CLPP*, adipose tissue (au) | -0.05 (-0.28 - 0.08) | -0.02 (-0.13 - 0.08) | 0.359 |
| *HSPD1*, adipose tissue (au) | 0.03 (-0.05 - 0.14) | -0.01 (-0.07 - 0.17) | 0.320 |
| *Satellite cells* | | | |
| *PAX7*, muscle (au) | -0.31 ± 0.30 | -0.26 ± 0.45 | 0.688 |
| PAX7 positive cells (PAX7+ cells/muscle fiber area) | -6.4 ± 3.6 | -3.9 ± 2.6 | 0.578 |
| *PAX7/MYOG* (ratio) | -0.63 ± 0.75 | -1.1 ± 0.22 | 0.500 |

Data are shown as mean ± SD (normally distributed variables), median (interquartile range, for skewed variables). P values were obtained using paired Wilcoxon rank-sum test to examine the differences on delta change (the change from baseline to 5 months) for anthropometric and clinical parameters between the leaner and heavier cotwins from the BMI-discordant pairs (n=16 twin pairs/32 individuals). NAAD, nicotinic acid adenine dinucleotide; NMN, nicotinamide mononucleotide; NAR, nicotinic acid riboside; Me4Py, N-methyl-4-pyridone-5-carboxamide; NADP, nicotinamide adenine dinucleotide phosphate; ADPR, ADP ribose; Au, arbitrary unit; *NRK1*, nicotinamide riboside kinase 1; *NAMPT,* nicotinamide phosphoribosyltransferase; *NMNAT3,* nicotinamide-nucleotide adenylyltransferase 3; *NAPRT,* nicotinic acid phosphoribosyltransferase; HbA1c, hemoglobin A1c; HOMA, homeostasis model assessment; AUC, area under curve; OGTT, oral glucose tolerance test; HDL, high-density lipoprotein; LDL, low-density lipoprotein; FFA, free fatty acids; BP, blood pressure; Hs-CRP, high sensitive C-reactive protein; 16S, 16 rRNA; CYTB, cytochrome b; DLOOP, D-loop region; *ERRα,* estrogen-related receptor α; *NRF1,* nuclear respiratory factor 1*; PGC1α, p*eroxisome proliferator-activated receptor gamma coactivator 1*α; TFAM*, transcription factor A; *MFN2,* mitofusin 2; *NDUFB8*, NADH:ubiquinone oxidoreductase subunit B8; *MT-ND5*, mitochondrially encoded NADH dehydrogenase 5; *SDHB*, succinate dehydrogenase complex iron sulfur subunit B; *UQCRC2*, ubiquinol-cytochrome c reductase core protein 2; *MT-CO1*, mitochondrially encoded cytochrome c oxidase 1; *ATP5A*, ATP synthase *α; CLPP*, caseinolytic mitochondrial matrix peptidase proteolytic subunit; *HSPD1*, heat shock protein family D (Hsp60) member 1; *CPT1β,* carnitine palmitoyltransferase 1B; *PAX7*, paired box 7 and *MYOG,* myogenin.

**Table S3. Glucose homeostasis in the twins from the BMI-discordant pairs upon NR supplementation.**

| Variable (unit) | All<br>Pre (*n*=32 individuals) | All<br>Post (*n*=32 individuals) | P Value | Leaner<br>Pre (*n*=16 individuals) | Leaner<br>Post (*n*=16 individuals) | P Value | Heavier<br>Pre (*n*=16) individuals | Heavier<br>Post (*n*=16 individuals) | P Value |
|---|---|---|---|---|---|---|---|---|---|
| HbA1c (mmol/mol) | 34 (32 - 36) | 35 (33 - 36) | 0.353 | 34 (32 - 35) | 35 (33 - 36) | 0.032 | 35 (34 - 36) | 35 (32 - 37) | 0.693 |
| Fasting glucose (mmol/l) | 5.6 (5.4 - 5.8) | 5.7 (5.5 - 5.9) | 0.027 | 5.6 (5.3 - 5.7) | 5.7 (5.3 - 5.8) | 0.483 | 5.7 (5.4 - 5.8) | 5.8 (5.6 - 5.9) | 0.018 |
| Fasting insulin (mlU/l) | 6.0 (4.5 - 9.8) | 7.7 (6.2 - 13.4) | 0.023 | 4.7 (3.8 - 6.7) | 7.0 (5.9 - 9.0) | 0.069 | 7.9 (5.3 - 12.5) | 12.1 (7.1 - 14.6) | 0.169 |
| Fasting C-peptide (nmol/l) | 0.51 (0.39 - 0.65) | 0.63 (0.50 - 0.77) | 0.002 | 0.47 (0.39 - 0.60) | 0.55 (0.48 - 0.65) | 0.022 | 0.56 (0.43 - 0.83) | 0.74 (0.59 - 0.84) | 0.036 |
| HOMA index | 1.5 (1.0 - 2.1) | 2.0 (1.5 - 3.3) | 0.016 | 1.1 (0.9 - 1.6) | 1.7 (1.4 - 2.4) | 0.073 | 2.0 (1.3 - 3.2) | 3.0 (1.7 - 3.8) | 0.135 |
| Matsuda index | 5.7 ± 2.5 | 4.4 ± 1.9 | <0.001 | 6.8 ± 2.4 | 5.3 ± 2.0 | 0.068 | 4.7 ± 2.1 | 3.6 ± 1.4 | 0.042 |
| Glucose AUC during OGTT | 15.7 ± 2.5 | 16.1 ± 2.7 | <0.001 | 15.2 ± 2.5 | 15.4 ± 2.6 | 0.286 | 16.2 ± 2.4 | 16.9 ± 2.8 | 0.208 |
| Insulin AUC during OGTT | 90 (72 - 135) | 116 (88 - 160) | <0.001 | 85 (68 - 117) | 112 (86 - 127) | 0.217 | 97 (78 - 175) | 154 (91 - 204) | 0.173 |
| Adiponectin (µg/ml) | 3.2 (2.1 - 4.3) | 3.2 (2.0 - 4.1) | 0.946 | 3.2 (2.7 - 3.8) | 3.3 (2.7 - 3.9) | 0.495 | 3.1 (1.6 - 5.2) | 2.8 (1.7 - 4.4) | 0.277 |

Data are shown as mean ± SD (normally distributed variables), median (interquartile range, for skewed variables). P values were obtained using paired Wilcoxon rank-sum test to examine the effects of NR supplementation on anthropometric and clinical parameters before and after NR intervention in the twins from the BMI-discordant pairs (n=16 twin pairs/32 individuals). HbA1c, hemoglobin A1c; HOMA, homeostasis model assessment; AUC, area under curve and OGTT, oral glucose tolerance test.

**Table S4. The effect of NR on cardiovascular parameters in the twins from BMI-discordant pairs.**

| Variable (unit) | All<br>Pre (*n*=32 individuals) | All<br>Post (*n*=32 individuals) | P Value | Leaner<br>Pre (*n*=16 individuals) | Leaner<br>Post (*n*=16 individuals) | P Value | Heavier<br>Pre (*n*=16 individuals) | Heavier<br>Post (*n*=16 individuals) | P Value |
|---|---|---|---|---|---|---|---|---|---|
| Cholesterol (total, mmol/l) | 4.7 ± 0.8 | 4.7 ± 0.8 | 0.346 | 4.7 ± 0.7 | 4.7 ± 0.7 | 0.568 | 4.7 ± 0.9 | 4.8 ± 1.0 | 0.470 |
| HDL (mmol/l) | 1.3 (1.2 - 1.6) | 1.2 (1.0 - 1.6) | 0.049 | 1.4 (1.2 - 1.5) | 1.3 (1.1 - 1.7) | 0.093 | 1.3 (1.1 - 1.7) | 1.2 (1.0 - 1.6) | 0.289 |
| LDL (mmol/l) | 3.0 ± 0.8 | 3.0 ± 0.5 | 0.723 | 3.0 ± 0.7 | 3.0 ± 0.5 | 0.752 | 3.1 ± 0.8 | 3.0 ± 0.5 | 0.887 |
| Triglycerides (mmol/l) | 1.0 (0.7 - 1.2) | 0.9 (0.8 - 1.3) | 0.046 | 0.81 (0.6 - 1.0) | 0.9 (0.7 - 1.1) | 0.313 | 1.0 (0.7 - 1.3) | 1.1 (0.8 - 1.4) | 0.088 |
| FFA (µmol/l) | 444 (376 – 551) | 431 (368 – 546) | 0.229 | 415 (362 - 500) | 392 (322 – 432) | 0.561 | 510 (428 - 700) | 483 (420 – 562) | 0.252 |
| Systolic BP (mmHg) | 133 ± 22 | 128 ± 19 | 0.531 | 131 ± 24 | 124 ± 19 | 0.683 | 135 ± 22 | 131 ± 19 | 0.753 |
| Diastolic BP (mmHg) | 83 ± 12 | 83 ± 11 | 0.875 | 83 ± 12 | 81 ± 11 | 0.889 | 84 ± 11 | 84 ± 12 | 1.000 |
| Pulse (per min) | 67 ± 11 | 67 ± 9 | 0.629 | 65 ± 9 | 66 ± 9 | 0.753 | 69 ± 12 | 69 ± 9 | 0.806 |
| Hs-CRP (mg/l) | 1.9 (0.7 - 3.9) | 1.6 (0.6 - 4.0) | 0.655 | 1.7 (0.6 - 3.0) | 1.1 (0.5 - 2.9) | 0.561 | 2.3 (0.7 - 5.8) | 2.5 (0.9 - 4.7) | 0.978 |
| Total homocysteine (µmol/l) | 11.0 (9.30 -12.0) | 12.0 (10.0 - 14.0) | 0.005 | 10.0 (9.55 -12.0) | 12.0 (10.5 - 14.0) | 0.003 | 11.0 (9.28 -12.8) | 11.5 (10.3 - 14.0) | 0.551 |

Data are shown as mean ± SD (normally distributed variables), median (interquartile range, for skewed variables). P values were obtained using paired Wilcoxon rank-sum test to examine the effect of NR supplementation on anthropometric and clinical parameters in the twins from the BMI-discordant pairs (n=16 twin pairs/32 individuals). HDL, high-density lipoprotein; LDL, low-density lipoprotein; FFA, free fatty acids; BP, blood pressure and Hs-CRP, high sensitive C-reactive protein.

**Table S10. The effect of 5-month placebo and NR treatment in the twins from the BMI-concordant pairs.**

| Variable (unit) | Placebo Pre (*n*=4 individuals) | Placebo Post (*n*=4 individuals) | P Value | Treated Pre (*n*=4 individuals) | Treated Post (*n*=4 individuals) | P Value |
|---|---|---|---|---|---|---|
| BMI (kg/m$^2$) | 31.5 ± 6.8 | 32.6 ± 7.3 | 0.125 | 32.0 ± 6.9 | 32.1 ± 6.7 | 0.875 |
| Weight (kg) | 87.2 ± 12.2 | 90.2 ± 13.2 | 0.125 | 89.1 ± 16.6 | 89.8 ± 15.2 | 0.581 |
| Waist-hip ratio | 0.9 ± 0.0 | 1.0 ± 0.0 | 0.250 | 0.9 ± 0.0 | 1.0 ± 0.1 | 0.250 |
| Body fat (%) | 34.1 ± 13.8 | 38.0 ± 12.3 | 0.250 | 33.2 ± 14.8 | 35.8 ± 13.3 | 0.750 |
| Visceral adipose tissue (cm$^3$) | 2017 ± 497 | 2275 ± 521 | 0.125 | 2217 ± 921 | 2171 ± 861 | 0.625 |
| Subcutaneous adipose tissue (cm$^3$) | 7297 ± 4799 | 7568 ± 4950 | 0.125 | 6996 ± 4831 | 6833 ± 4751 | 0.375 |
| Adipocyte cell number (per billion) | 233 (197 - 308) | 289 (235 - 325) | 1.000 | 168 (147 - 204) | 315 (246 - 396) | 0.125 |
| Adipocyte cell diameter (µM) | 89 ± 11 | 99 ± 8.6 | 0.375 | 96 ± 12 | 90 ± 5.6 | 0.125 |
| Adipocyte cell volume (pL) | 500 ± 154 | 734 ± 161 | 0.250 | 617 ± 219 | 573 ± 123 | 0.625 |
| Adipocyte cell weight (ng) | 407 (364 - 500) | 702 (589 - 785) | 0.250 | 584 (427 - 722) | 559 (495 - 587) | 0.625 |
| Liver fat (%) | 5.2 (0.87 - 9.9) | 6.5 (1.5 - 12.0) | 0.181 | 3.4 (2.0 - 5.4) | 3.1 (1.9 - 4.8) | 0.125 |
| Lean tissue mass (kg) | 56.2 ± 5.5 | 53.0 ± 8.8 | 0.750 | 58.8 ± 4.9 | 54.3 ± 9.7 | 1.000 |
| Total bone mass (kg) | 3.2 ± 0.6 | 3.0 ± 0.7 | 0.500 | 3.3 ± 0.5 | 3.0 ± 0.7 | 0.250 |
| Basal metabolic rate (kcal/day) | 1782 ± 266 | 1813 ± 260 | 0.250 | 1814 ± 298 | 1842 ± 262 | 0.125 |
| Caloric intake (kcal/day) | 1872 ± 419 | 1423 ± 730 | 0.250 | 2134 ± 883 | 1831 ± 1120 | 0.250 |
| Protein intake (g) | 81.5 (63.2 - 100.2) | 89.5 (61.6 - 91.8) | 1.000 | 84.0 (65.7 - 110.1) | 69.2 (47.3 - 89.9) | 0.125 |
| Fat intake (g) | 69.8 ± 17.5 | 52.7 ± 29.8 | 0.250 | 87.1 ± 39.5 | 70.2 ± 53.2 | 0.250 |
| Carbohydrate intake (g) | 203.5 ± 44.4 | 134.9 ± 66.8 | 0.500 | 221.2 ± 93.2 | 187.0 ± 131.5 | 0.625 |
| Niacin equivalent (mg/day) | 32.5 (30.6 - 36.5) | 40.9 (29.1 - 44.1) | 1.000 | 35.8 (31.9 - 41.6) | 29.9 (21.7 - 37.7) | 0.125 |
| Physical activity (Total Baecke) | 8.1 ± 0.7 | 8.3 ± 1.5 | 0.500 | 8.0 ± 1.7 | 8.0 ± 1.5 | 1.000 |
| Alcohol intake (doses/week) | 1.0 (0.5 - 1.4) | 0.0 (0.0 – 7.0) | 1.000 | 2.0 (0.0 - 4.6) | 0.0 (0.0 - 2.5) | 1.000 |
| Current smoker (number) | 2 | 1 | - | 1 | 1 | - |
| HbA1c (mmol/mol) | 34.5 (33.0 – 36.3) | 35.5 (34.5 – 36.3) | 1.000 | 36.5 (35.0 – 37.8) | 37.0 (35.5. – 38.5) | 0.371 |
| Fasting glucose (mmol/l) | 5.4 (5.0 - 5.8) | 5.4 (5.2 - 5.6) | 0.850 | 5.8 (5.6 - 5.4) | 5.4 (5.2 - 5.5) | 0.625 |
| Fasting insulin (mlU/l) | 7.7 (3.9 - 11.5) | 6.1 (.3 - 11.9) | 0.875 | 9.7 (4.1 - 14.9) | 10.4 (4.4 - 16.2) | 0.125 |
| Fasting C-peptide (nmol/l) | 0.7 (0.4 – 1.0) | 0.6 (0.4 - 1.0) | 0.875 | 0.8 (0.4 - 1.2) | 0.8 (0.4 - 1.1) | 0.250 |
| HOMA index | 1.8 (1.0 - 2.7) | 1.5 (0.8 - 2.8) | 0.875 | 2.5 (1.0 - 3.8) | 2.4 (1.0 - 4.0) | 1.000 |
| Matsuda index | 8.8 ± 6.9 | 6.9 ± 5.0 | 0.250 | 7.5 ± 6.5 | 5.3 ± 4.5 | 0.125 |
| Glucose AUC during OGTT | 14.8 ± 4.1 | 15.4 ± 3.2 | 0.875 | 16.8 ± 3.3 | 24.4 ± 13.1 | 0.625 |
| Insulin AUC during OGTT | 90 (30 - 161) | 107 (49 - 190) | 0.125 | 115 (39 - 188) | 108 (45 - 200) | 0.625 |
| Adiponectin (µg/ml) | 3.4 (2.8 - 6.8) | 3.2 (2.4 - 6.9) | 0.750 | 5.2 (3.6 - 6.9) | 5.0 (3.7 - 6.2) | 1.000 |
| Cholesterol (total, mmol/l) | 4.7 ± 0.5 | 4.9 ± 0.5 | 0.181 | 4.8 ± 0.8 | 4.7 ± 0.5 | 0.371 |
| HDL (mmol/l) | 1.3 (1.2 - 1.4) | 1.4 (1.3 - 1.4) | 0.875 | 1.4 (1.2 - 1.6) | 1.3 (1.1 - 1.4) | 0.098 |

| | | | | | | |
|---|---|---|---|---|---|---|
| LDL (mmol/l) | 3.1 ± 0.41 | 3.4 ± 0.57 | 0.174 | 3.3 ± 0.45 | 3.4 ± 0.47 | 0.174 |
| Triglycerides (mmol/l) | 1.0 (0.9 - 1.0) | 1.0 (0.8 - 1.3) | 0.875 | 0.9 (0.7 - 1.2) | 0.9 (0.8 - 1.1) | 1.000 |
| FFA (μmol/l) | 427.7 (325.4 - 531.4) | 415.3 (347.7 - 485.2) | 0.625 | 394.1 (332.1 - 481.9) | 456.3 (346.6 - 551.1) | 0.625 |
| Systolic BP (mmHg) | 114 ± 12 | 122 ± 11 | 0.250 | 126 ± 8 | 129 ± 16 | 0.625 |
| Diastolic BP (mmHg) | 74 ± 9 | 80 ± 12 | 0.125 | 80 ± 5 | 82 ± 13 | 0.713 |
| Pulse (per min) | 63 ± 8 | 63 ± 13 | 1.000 | 61 ± 5 | 61 ± 13 | 0.875 |
| Hs-CRP (mg/l) | 1.6 (1.3 - 11) | 2.0 (0.9 - 5.5) | 0.250 | 1.0 (0.6 - 3.3) | 0.4 (0.4 - 5.1) | 1.000 |
| Total homocysteine (μmol/l) | 13.0 (12.2 - 13.8) | 14.0 (12.1 - 15.8) | 0.414 | 12.5 (11.5 - 15.0) | 13.5 (12.1 - 17.0) | 0.198 |
| ALT (U/l) | 24 (18 - 39) | 24 (21 - 33) | 1.000 | 23 (22 - 28) | 27 (21 - 34) | 1.000 |
| AST (U/l) | 27 (25 - 30) | 23 (22 - 26) | 0.250 | 27 (24 - 30) | 31 (26 - 34) | 0.581 |
| Creatinine (μmol/l) | 74 ± 13 | 75 ± 8 | 0.875 | 79 ± 12 | 76 ± 5 | 0.625 |
| Haemoglobin (g/l) | 137 ± 10 | 132 ± 6 | 0.197 | 136 ± 13 | 130 ± 12 | 0.197 |
| Haematocrit (%) | 40.8 ± 2.9 | 39.5 ± 2.1 | 0.173 | 40.5 ± 3.9 | 38.5 ± 3.0 | 0.174 |
| Erythrocytes ($x10^{12}$/l) | 4.5 ± 0.4 | 4.5 ± 0.3 | 0.875 | 4.6 ± 0.5 | 4.5 ± 0.4 | 0.250 |
| MCV (fl) | 90.3 ± 2.1 | 88.3 ± 3.9 | 0.250 | 87.8 ± 4.2 | 86.3 ± 5.6 | 0.346 |
| MCH (pg) | 30.5 ± 1.0 | 29.8 ± 0.96 | 0.149 | 29.8 ± 1.7 | 29.0 ± 2.2 | 0.371 |
| MCHC (g/l) | 338.0 (335.5 - 339.8) | 335.0 (332.5 - 338.0) | 1.000 | 338.5 (334.5 - 342.5) | 335.5 (331.3 - 339.3) | 0.625 |
| Leukocytes ($x10^9$/l) | 7.3 ± 3.0 | 7.0 ± 2.4 | 0.875 | 6.2 ± 1.7 | 6.0 ± 0.9 | 1.000 |
| Thrombocytes ($x10^9$/l) | 234 ± 29 | 260 ± 48 | 0.197 | 260 ± 53 | 238 ± 31 | 0.375 |
| fP-Fe (μmol/l) | 20.0 ± 6.4 | 20.4 ± 3.2 | 1.000 | 20.0 ± 9.1 | 16.6 ± 5.1 | 0.875 |

Data are shown as mean ± standard deviation or median (interquartile range). P values were obtained using paired Wilcoxon rank-sum test to examine the effect of placebo and NR treatments on anthropometric and clinical parameters in the twins from the BMI-concordant pairs (n= 4 pairs/8 individuals). HbA1c, hemoglobin A1c; HOMA, homeostasis model assessment; AUC, area under curve; OGTT, oral glucose tolerance test; HDL, high-density lipoprotein; LDL, low-density lipoprotein; FFA, free fatty acids; BP, blood pressure; Hs-CRP, high sensitive C-reactive protein; ALT, alanine aminotransferase; AST, aspartate transaminase; MCV, mean corpuscular volume; MCH, mean corpuscular haemoglobin; MCHC, mean corpuscular haemoglobin concentration; fP, fasting plasma and Fe, iron.

**Table S11. The effect of NR on safety parameters in the twins from BMI-discordant pairs.**

| Variable (unit) | All<br>Pre (*n*=32 individuals) | All<br>Post (*n*=32 individuals) | P Value | Leaner<br>Pre (*n*=16 individuals) | Leaner<br>Post (*n*=16 individuals) | P Value | Heavier<br>Pre (*n*=16 individuals) | Heavier<br>Post (*n*=16 individuals) | P Value |
|---|---|---|---|---|---|---|---|---|---|
| ALT (U/l) | 22 (18 - 43) | 26 (17 - 43) | 0.674 | 20 (17 - 34) | 26 (19 - 36) | 0.041 | 33 (20 - 49) | 27 (17 - 44) | 0.155 |
| AST (U/l) | 24 (22 - 31) | 25 (21 - 30) | 0.626 | 24 (20 - 29) | 24 (21 - 35) | 0.081 | 24 (23 - 31) | 25 (22 - 28) | 0.293 |
| Creatinine (μmol/l) | 74 ± 13 | 74 ± 12 | 0.650 | 73 ± 14 | 75 ± 12 | 0.551 | 74 ± 13 | 74 ± 12 | 0.924 |
| Haemoglobin (g/l) | 142 ± 9 | 140 ± 9 | 0.043 | 142 ± 8 | 140 ± 8 | 0.312 | 143 ± 9 | 141 ± 10 | 0.073 |
| Haematocrit (%) | 42.0 ± 2.2 | 42.1 ± 2.2 | 0.641 | 41.9 ± 2.1 | 42.4 ± 1.9 | 0.212 | 42.1 ± 2.4 | 41.9 ± 2.6 | 0.668 |
| Erythrocytes (x$10^{12}$/l) | 4.8 ± 0.3 | 4.8 ± 0.3 | 0.991 | 4.8 ± 0.37 | 4.8 ± 0.32 | 0.244 | 4.8 ± 0.31 | 4.8 ± 0.31 | 0.268 |
| MCV (fl) | 87.9 ± 3.5 | 88.0 ± 3.3 | 0.829 | 88.4 ± 4.2 | 88.3 ± 3.7 | 1.000 | 87.4 ± 2.6 | 87.7 ± 2.9 | 0.681 |
| MCH (pg) | 29.8 ± 1.3 | 29.4 ± 1.4 | 0.013 | 29.9 ± 1.5 | 29.3 ± 1.7 | 0.009 | 29.6 ± 1.2 | 29.6 ± 1.1 | 0.766 |
| MCHC (g/l) | 338 (334 - 342) | 333 (330 - 340) | 0.005 | 338 (334 - 342) | 332 (327 - 338) | 0.010 | 337 (335 - 342) | 336 (331 - 341) | 0.300 |
| Leukocytes (x$10^{9}$/l) | 6.4 ± 1.5 | 6.3 ± 1.8 | 0.134 | 6.5 ± 1.8 | 6.3 ± 2.0 | 0.277 | 6.3 ± 1.2 | 6.3 ± 1.7 | 0.244 |
| Thrombocytes (x$10^{9}$/l) | 243 ± 55 | 248 ± 62 | 0.199 | 244 ± 60 | 252 ± 72 | 0.301 | 241 ± 52 | 244 ± 52 | 0.569 |
| fP-Fe (μmol/l) | 19.0 ± 6.9 | 16.7 ± 5.9 | 0.078 | 19.8 ± 7.6 | 16.6 ± 5.9 | 0.084 | 18.1 ± 6.3 | 16.7 ± 6.1 | 0.349 |

Data are shown as mean ± standard deviation or median (interquartile range). P values were obtained using paired Wilcoxon rank-sum test to examine the effect of NR supplementation on safety measures in the twins from the BMI-discordant pairs (n=16 twin pairs/32 individuals). ALT, alanine aminotransferase; AST, aspartate transaminase; MCV, mean corpuscular volume; MCH, mean corpuscular haemoglobin; MCHC, mean corpuscular haemoglobin concentration; fP, fasting plasma and Fe, iron.
